# IBS stress reactivity phenotype is associated with blood transcriptome profiles and microstructural and functional brain changes

**DOI:** 10.1101/2024.08.07.24311369

**Authors:** Jennifer S Labus, Desiree R. Delgadillo, Steve Cole, Chencai Wang, Bruce Naliboff, Lin Chang, Benjamin M. Ellingson, Emeran A. Mayer

## Abstract

**Background & Aims:** Clinical evidence suggests significant interindividual differences in stress reactivity (SR), but biological mechanisms and therapeutic implications of these differences are poorly understood. We aimed to identify the biological basis of increased SR by investigating associations between a psychometric-based phenotype with blood transcriptomics profiles of increased sympathetic nervous system (SNS) activation and brain imaging phenotypes in irritable bowel syndrome (IBS) participants and healthy controls (HCs).

**Methods:** A cross-sectional observational study design, transcriptomics profiling, multimodal brain imaging, and psychosocial assessments were obtained in 291 female and male IBS participants and HCs. Prior to analyses, unsupervised clustering was applied to derive high and low SR subgroups across participants based on two measures of SR. General linear models tested for SR group differences in clinical and biological parameters. Exploratory analyses examined associations between SR group-specific brain alterations and gene expression.

**Results:** The high, compared to low SR group showed greater cyclic AMP response element-binding protein (CREB) gene expression consistent with tonic SNS activity and proinflammatory changes in whole blood. Brain imaging showed neuroplastic changes in the high SR group consistent with an upregulation of ascending arousal systems and sensory processing and integration regions, and functional connectivity changes in the central autonomic network. SR moderated the sex difference in extraintestinal symptoms.

**Conclusions:** The findings support a model of tonically increased SNS activity as a plausible risk factor for increased autonomic reactivity to psychosocial stressors and low grade immune activation in both IBS and HCs, with a greater prevalence in IBS. These findings may have important implications for personalized treatment interventions in IBS.

## Introduction

Irritable bowel syndrome (IBS) is a stress-sensitive disorder of brain gut interactions, currently defined by symptom criteria of chronically recurring abdominal pain associated with altered bowel habits.^1^ A wealth of information has demonstrated evidence to support important roles of visceral hypersensitivity and of altered autonomic nervous system modulation of gut functions in the pathophysiology of IBS, even though the role and underlying mechanisms of altered stress responsiveness has not been studied as a risk factor and disease modifier.

Chronic stress is associated with an increased odds of having IBS and the persistence of symptoms.^2^ Seventy percent of patients with IBS have a history of early life and adulthood stressful life events^3–6^ and reduced resilience^7^ (adapting positively to stress) compared to healthy controls (HCs). Furthermore, psychosocial stress and food intake are considered the major factors underlying symptom flares, and are associated with more severe symptoms and worse health-related quality of life.^8–10^ The acute stress response system is composed of the sympathetic nervous system (SNS) and the hypothalamic pituitary adrenal (HPA) axis, which are coordinated by the central autonomic network, including several brainstem nuclei. The physiological reaction to stressful life events and the subjective perception of stress shows significant inter-individual variations which are likely to influence the various clinical presentation of functional and inflammatory GI disorders^11^ which may have significant implications for individualized treatment strategies.

Heightened stress responsivity (SR) refers to an exaggerated or heightened physiological or emotional response to stress, a condition where individuals respond more intensely to stressors compared to the average person. This heightened reactivity can manifest in various ways, including physical symptoms (rapid heart rate, elevated blood pressure,^12^ increased sigmoid contractions, increased visceral sensitivity, excessive sweating, and other physical signs of stress), emotional and psychological responses (intense feelings of anxiety, irritability, or depression in response to stressful situations) and behavioral changes (overreaction to minor stressors, difficulty in calming down after stress, or avoidance of situations that are perceived to be stressful). Evidence suggests, that individuals with increased SR have a higher prevalence and severity of several stress-related disorders, including IBS, and increased SR in healthy people has been suggested as a risk factor for developing IBS and other often comorbid disorders.^13^ It is also possible that sex differences in SR may play a role in the increased sensory sensitivity and higher rates of comorbidity found in females compared to males for IBS,^14,15^ as well as other chronic pain conditions such as urological chronic pelvic pain syndrome,^16^ but to date there has been little study of the biological underpinnings of these differences. Here we investigate the biological mechanisms underlying increased SR, the effects of increased SR on reports of comorbidity, and how SR may moderate reported sex differences in IBS.

The overall goal of this study was to identify the biological basis of increased SR in a large population of IBS and HCs by studying associations between a psychometric--based phenotype of increased SR with several peripheral and central biomarkers. We aimed to test the main hypothesis that a subgroup of individuals with IBS as well HCs, with high SR identified based on clinical characteristics, will show evidence for a biological phenotype of increased transcription of SNS- and inflammation-related genes, as well as functional and microstructural brain changes that may underlie development and persistence of stress related disorders. We also examine the clinical correlates of this SR phenotype within IBS and HC in order to test whether even in HCs, heightended SR may be a vulnerability factor for chronic pain.

Our findings confirmed the main hypothesis and demonstrate that the High SR phenotype is characterized by a history of early life adversity, lower resilience, higher psychosocial comorbidity and greater intestinal and extraintestinal symptom severity, and that some of the observed brain changes are associated with SNS-related gene expression. Furthermore, even though there are no sex-related differences in the prevalence of the High SR phenotype, SR moderates the sex differences in comorbidity in IBS.

## Methods

### Subject Population

Male and female HCs and IBS participants were prospectively enrolled in a cross-sectional study involving observational MRI brain imaging and evaluation of IBS symptoms based on ROME III^17^ and IV^14^ criteria from 2013-2019. A gastroenterology (GI) clinician obtained histories and physical examinations and confirmed the diagnosis of IBS. All procedures complied with the principles of the Declaration of Helsinki and were approved by the Institutional Review Board.

Exclusionary criteria for all participants included pregnancy or lactation, tobacco dependence (smoked half a package of cigarettes or more daily), substance abuse, abdominal surgery other than appendectomy or cholecystectomy, current or past psychiatric illness, extreme strenuous exercise, major medical or neurological conditions, and current regular use of analgesic drugs (including narcotics, α2-δ ligands, and opioids).

Medication usage such as antidepressants (including selective serotonin uptake inhibitors, nonselective serotonin reuptake inhibitors, low-dose tricyclic anti-depressants) was permitted only if participants were on a stable dose for a minimum of 3 months. Participants were not excluded for NSAIDS usage such as diclofenac. However, they were asked to refrain from taking this medication 12 hours prior to their brain imaging visit and blood sample. All female participants were premenopausal confirmed by self-report and were scanned during the follicular phase of the menstrual cycle based on last menstrual period and progesterone level.

### Assessment of Subjective Parameters

#### Psychosocial phenotyping

Self-reported ongoing stress was measured using Cohen’s Perceived Stress Scale (PSS).^18^ Emotional reactivity was assessed with the International Personality Item Pool-Neuroticism (IPIP-N).^19^ Current anxiety symptoms were assessed with the Hospital Anxiety and Depression scale.^20^ Trait anxiety was measure used the State Trait Anxiety Inventory.^21^ GI-specific anxiety was assessed using the Visceral Sensitivity Index.^22,23^ Pain Catastrophizing was evaluated with the catastrophizing scale of the Coping Strategies Questionnaire.^24^

Early adverse life experiences were assessed using the Adverse Childhood Events questionnaire (ACE) and the Early Traumatic Inventory–Self Report (ETI-SR). The ACE captures emotional, physical, and sexual abuse; parents who were divorced or separated, treated violently; and household member with substance abuse, with mental illness, and who was incarcerated.^25^ The ETI-SR was used to assess histories of childhood traumatic and adverse life events that occurred before the age of 18 years old and covers four domains: general trauma, physical, emotional, and sexual abuse.^26^

Different aspects of perceived resilience were measured using The Connor-Davidson Resilience Scale (CDRISC) and Brief Resilience Scale (BRS) which differ in complexity.^27–29^ The CDRISC is focused on internal and external resources that aid in recovery from stress and has 5 domains: persistence, emotional-cognitive, adaptability, control-meaning, and meaning. The total CDRISC score ranges 0-100, with higher scores indicating higher resilience. The BRS assess the ability to “bounce back” from stress and adversity. Higher scores on the BRS reflect higher resilience in terms of bounce-back ability.

#### IBS Symptom measures

IBS symptom severity was measured using the IBS Severity Scoring System (IBS-SSS) which assesses abdominal pain intensity and frequency, distention, dissatisfaction with bowel habits, and impact of IBS on quality of life over the past 10 days.^30^ Disease-specific quality of life (QOL) in the past month was evaluated using the total score from the IBS-QOL which assesses the interference of IBS symptoms on various aspects of daily activities.^31^

#### Extraintestinal symptom assessment

Extraintestinal symptom prevalence was assessed using several assessment tools. Somatic symptom severity was measured with the Patient Health Questionnaire without GI items (PHQ-12).^32^ The Pennebaker Inventory of Limbic Languidness (PILL) used to assess enhanced interoception and the tendency to notice and report a broad array of physical symptoms and sensations.^33^ The Complex Medical Symptom Inventory (CMSI) was used to delineate the presence of common somatic symptoms of discomfort or pain as well as sensory sensitivity to nonpainful environmental stimuli over the lifetime (CMSI_Lftm) and for 3 months out of the past year (CMSI_12m).^34^ Scores on the somatic awareness (CMSI_SA) and sensory sensitivity(CMSI_SS) subscales were computed.^35^

#### Assessment of biological parameters

Details on Blood RNA profiling and MRI acquisition protocols, preprocessing and computational statistics can be found in Supplemental Methods.

### Statistical analysis of subjective and biological parameters

#### Unsupervised learning to subtype participants by Stress Reactivity

SR groups were defined by the PSS (perceived ongoing stress) and IPIP-N (emotional reactivity to stressors) using unsupervised learning across HC and IBS participants.^11,36^ This subtyping was performed in the two study cohorts providing data for the blood transcriptome analyses and brain imaging. Specifically, partitioning around medoids (PAM) using Manhattan’s distances was used to derive SR subgroups based on these two baseline measures of SR. The optimal number of clusters was selected using validity indices calculated using the R library NBclust.

#### RNA profile analysis

Transcript abundance values were normalized to 11 standard reference genes^37^ and log2-transformed for linear model analyses quantifying differences in gene expression as a function of High-vs. Low-SR group while controlling for age, sex, race/ethnicity, education, income, and mRNA transcripts indicating relative prevalence of major leukocyte subsets (*CD3D, CD4, CD8A, CD19, NCAM1*/CD56, *FCGR3A*/CD16, *CD14*). As in previous research,^38^ primary mixed effect linear model analyses examined 16 pro-inflammatory gene transcripts (identified from a set of 19 pre-specified indicators used in previous research, after removal of 5 showing minimal expression variance in this sample; SD < .5 log_2_ units) with genes treated as repeated measures. Secondary analyses used TELiS promoter-based bioinformatics analyses to analyze all genes (transcriptome-wide) showing > 50% differential expression in high-vs. low-stress groups (again controlling for covariates above), testing for differential prevalence of transcription factor-binding motifs (TFBMs) in gene core promoter sequences for key pro-inflammatory transcription factors (NF-κB and AP-1, assessed by TRANSFAC position-specific weight matrices V$AP1$_Q2_01 and V$CREL_01), the cyclic AMP response element-binding protein (CREB) family of factors involved in beta-adrenergic signaling from the sympathetic nervous system (V$CREB_Q3), and the glucocorticoid receptor (GR; V$GR_Q6) involved in cortisol signaling from the hypothalamus-pituitary-adrenal axis.^39^ Analyses were conducted in 9 combinations of core promoter length (−300, −600, and −1000 to +200 bp relative to the RefSeq transcription start site) and detection stringency (TRANSFAC mat_sim = .80, .90, and .95), with log_2_-transformed ratios of binding site prevalence in up-regulated vs. down-regulated genes averaged across the 9 parametric combinations and tested for statistically significant deviation from the null hypothesis of 0 (1-fold) using bootstrap standard errors controlling for correlation among genes as previously described.^38^ In an exploratory analysis, we also computed conserved transcriptional response to adversity (CTRA), pro-inflammatory, and anti-viral gene expression composite scores 19 inflammatory genes and 32 antiviral genes involved in Type I interferon responses and antibody production.^40^

#### DTI analyses

Using AFNI (Analysis of Functional NeuroImages),^41^ voxel-wise differences in DTI measurements were evaluated in the brain and brainstem using a general linear model.. Differences between the High and Low SR groups in the fractional Anisotropy (FA) and Median Diffusivity (MD), age, sex, BMI and diagnosis were included as covariates. FA and MD are standard measures of fiber density/connectivity for involved white matter regions, and measures of higher cellularity/cell packing for gray matter areas. For whole brain analysis, of cortical and subcortical white matter a cluster threshold was applied from permutation tests by estimating data’s smoothness through command 3dFWHMx in AFNI, and then estimating the cluster extent thresholds at a level of significance, p < 0.05, through command 3dClustSim in AFNI.^15^ Brainstem regions of interest (ROIs) were based on the Harvard Ascending Arousal Network (AAN) Atlas: locus coeruleus (LC), dorsal (DRN) and median raphe (MRN) nuclei, parabrachial complex (PBC), vental tegmental area (VTA), midbrain reticular formation (MRF)and the periaqueductal gray (PAG).^42^ For brain ROI interest analysis, a binary image with 1 assigned to pixels containing the region of interest and 0 assigned to regions outside prior to analysis was applied to mask out the regions of non-interest for analysis.

#### Resting state analysis

Using R, contrasts analysis was applied using the general linear model to test for differences in the resting strength of the regions of interest in the central autonomic network (CAN) and the brainstem nuclei (See **Supplemental Methods)**. We used p<.05 as a reporting threshold and also correct for the number of regions tested using a false discovery rate (FDR). Additionally, we provide a measure of the effect size difference, Cohen’s d where values are interpreted as low (d = 0.20), moderate (d = 0.50), and high (d = 0.80).

#### Association analyses

To investigate the link between DTI brainstem nuclei and RS ROIs showing differences between the SR groups and gene expression composite scores and the expression of the individual genes in the set, we employed general linear models controlling for diagnosis, sex, age, BMI, race (white yes/no), and education. We applied FDR correction for the number of regions tested and report standardized beta(Std B) values which are a measure of effect size and are interpreted 0.10–0.29 small, 0.30–0.49 medium, and 0.50 large.^43^

Independent sample t-tests were used to characterize differences between the SR subgroups (Low-High SR) across and within diagnosis (HC, IBS) on demographic and psychosocial variables. Within IBS participants, independent t-test were also applied to determine SR differences in IBS symptoms and comorbidity measures. Differences in frequencies were assessed via two-tail Fisher Exact Tests. Effect size differences were estimated using Cohen’s d and 95% Confidence intervals. Additionally, moderator analyses was applied to investigate whether sex differences in the comorbidity assessments in IBS were moderated by SR using the general linear model. Here we modeled the main effects for SR group and sex along with their interaction. A significant interaction effect suggests moderation.

## Results

### Study cohorts

The entire study sample consisted of 291participants (194 females) including 181 participants who met Rome III or Rome IV symptom criteria depending on year of enrollment for IBS (128 females) and 110 healthy controls (HCs; 66 females). Of these 253 participants had brain imaging data (84 male and 169 female (n=169) subjects, including 152 participants who met Rome III symptom criteria for IBS (107 females) and 101 healthy controls (HCs; 62 females). A total of 120 participants provided samples for blood transcriptome profile analysis including 82 IBS (60 females) and 38 HCs (25 females). A total of 82 subjects had paired imaging and blood transcriptome profiles.

### SR subgrouping

Unsupervised learning was applied to two study cohorts, those providing samples for blood transcriptomics analyses (N=120) and those with brain imaging (N=253), to subtype participants into High and Low SR group. For the 120 participants providing blood samples, 64 were clustered as Low SR (45 females[55%], 38 IBS[59%]) and 56 as high SR (38 females[68%], 44 IBS[76%], (see **Supplemental Figure 1**). In this cohort, there was no statistically significant difference between the proportion of males and females comprising the SR Groups (Fisher’s exact test, OR=0.89,p=0.84). IBS participants were twice as likely to be in the High SR group compared to the HC group (OR= 2.5, p=.03).

Next, clustering analyses was applied to the 253 HC (N=101; 62 females [61.4%]) and IBS (N=152; 107 females [70.4%]) participants with brain imaging data. Again, the algorithm identified two clusters subtyping participants into High and Low SR groups. As with the study cohort with blood sample only, the clusters did not show distinct separation (see **Supplemental Figure 2**). This was largely due to overlap between the two groups on the IPIP-N (see **Supplemental Figure 3**). To create a cleaner more distinct phenotypic grouping, we further refined cluster membership by removing subjects with overlapping IPIP scores, Low Stress (IPIP<=20) and High Stress (IPIP>29). This final sample was comprised by 168 participants (70 HC, 98 IBS) belonging to High SR (N=68, 49 females [72%], 55 IBS [56%], 13 HC [18.6%]) and Low SR (N=100, 65 females [65%], 43 IBS [43%]) groups. Of these 168 participants, a total of 50 participants had both blood transcriptome data and imaging data including 34 Low SR (23 females, 17 IBS) and 16 High SR (12 females, 14 IBS). As observed previously, there were no differences between the proportion of males and females comprising the SR Groups (Fisher’s exact test, OR=1.39,p=0.40), while IBS participants compared to HCs were more likely to be in the High SR group (OR=5.54.p=2.2.e-16). There were no significant differences in age or proportion of males and females comprising the IBS and HC groups

### Description of High and Low SR Groups for imaging cohort (N=168)

### SR differences on demographic and psychosocial assessments

#### Differences between the high and low SR groups across diagnosis

As shown in **Table 1**, the High SR group (combined IBS and HCs participants) had significantly higher state anxiety (Cohen’s d = −2.31, p = 7.59e^-32^), trait anxiety (d = −1.63, p = 1.12 e^-19^), and VSI scores (d = −0.84, p = 2.98e^-7^). The High SR group also reported significantly greater overall ETI-SR scores (d = −.067, p = 3.91 e^-5^) and also reported more early adverse life experiences as measured by the ACE total score (d = −0.61, p = 1.43e^-4^. The High SR showed less resilience as measured by the BRS (d = 1.54, p = 4.98e^-18^) and the CD-RISC subscale scores (see Table 1). No differences were observed between the proportion of men and women in the High and Low SR groups (Fisher exact test, *p* = 0.17). Means and SDs for all comparisons are provided in **Supplemental Table 1**.

**Table 1.** Across diagnosis comparison of low versus high SR groups on demographic and psychosocial variables.

| Variables | t | df | P | Cohen's d (95% CIs) |
| --- | --- | --- | --- | --- |
| ACE Total Score | -3.89 | 166 | $1.43 \times 10^{-4}$ | -0.61 (-0.93- -0.30) |
| Age | 1.17 | 166 | 0.24 | 0.18 (-0.13-0.49) |
| BMI | 2.12 | 166 | $3.54 \times 10^{-2}$ | 0.33 (0.023-0.64) |
| BRS Score | 9.75 | 165 | $4.98 \times 10^{-18}$ | 1.54 (1.19- 1.89) |
| CDRISC Adaptability/Ability to Bounce Back | 9.65 | 166 | $9.05 \times 10^{-18}$ | 1.52 (1.17-1.86) |
| CDRISC Emotional/Cognitive Control Under Pressure | 7.46 | 166 | $4.65 \times 10^{-12}$ | 1.17 (0.84-1.50) |
| CDRISC Meaning/Spiritual | 4.98 | 164 | $1.61 \times 10^{-6}$ | 0.79 (0.46-1.11) |
| CDRISC Persistence/Tenacity and Self-efficacy | 10.07 | 166 | $6.35 \times 10^{-19}$ | 1.58 (1.23-1.93) |
| CDRISC Meaning/Control | 10.60 | 165 | $2.33 \times 10^{-20}$ | 1.67 (1.31-2.03) |
| CSQ Catastrophizing Score | -5.68 | 161 | $6.21 \times 10^{-8}$ | -0.91 (-1.23- -.58) |
| Early Trauma Inventory - Total Score | -4.23 | 164 | $3.91 \times 10^{-5}$ | -0.67 (-0.99- -0.35) |
| HAD Anxiety | -10.35 | 166 | $1.12 \times 10^{-19}$ | -1.63 (-1.98- -1.27) |
| IPIP Neuroticism | -33.23 | 166 | $2.99 \times 10^{-75}$ | -5.22 (-5.86- -4.58) |
| PSS Score | -11.57 | 166 | $4.42 \times 10^{-23}$ | -1.82 (-2.18- -1.45) |
| STAIT Anxiety | -14.69 | 166 | $7.59 \times 10^{-32}$ | -2.31 (-2.70- -1.91) |
| VSI Score | -5.34 | 166 | $2.98 \times 10^{-7}$ | -0.84 (-1.16- -0.52) |

SR, stress reactivity; ACE, Adverse Childhood Experience survey; BMI, body mass index; BRS, Brief Resilience Scale; CDRISC, Conner-Davidson Resilience Scale; CSQ, Coping Strategies Questionnaire; HAD, Hospital Anxiety and Depression Scale; IPIP, International Personality Item Pool; PSS, perceived stress scale; STAI-T, Spielberger State Trait Anxiety Inventory-Trait; VSI, Visceral Sensitivity Index.

#### SR group differences within IBS or HC (see Supplemental Table 2)

Examining within IBS and HC groups, the VSI scores (GI-symptom related anxiety) only showed differences between High and Low SR in HC (d = - 0.75, *p* = 0.02 but not within IBS (d = −0.25, *p* = 0.23). Total ETI-SR score (early life trauma) was significantly higher in the High compared to Low SR group within HC (d = −0.85, *p* = 0.01) and IBS (d = −.44, *p* = 0.03 (See Supplemental Table 2).

### SR differences in IBS symptoms

Examining the IBS symptom-related measures, IBS participants in the High SR group, compared to the Low SR group, showed significantly higher IBS symptom severity as assessed by increased IBS-SSS scores (Cohen’s d = −0.54, *p* = 0.01) and reduced IBS-QOL (Cohen’s d = .44, *p* = .04). (see **Supplemental Table 3 and Supplemental Table 4**.)

### Influence of SR on Comorbidity measures in IBS

With in the IBS Cohort, High SR compared to low SR had significantly higher levels of comorbidity as measured by the PILL, d=-.90, p=2.7E-05, PHQ-12 score, d=-0.55, p= 7.9E-03, CMSI_SA, d=-0.65, 2.3E-030, CMSI_12mo score=-0.64, p= 2.9E-03 (see **Supplemental Tables 3 and 4**).

Additionally, SR was a moderator of sex difference in the CMSI_SA(p=3.21e-2), CMSI_Lftm(p=4.41e-2), CMSI_12m(p=4.54e-2) and the PHQ(5.46e-2). Namely, sex differences in these comorbidity measures were only observed in the High SR group (see **Supplemental Figure 4**). Examining the within group SR group contrasts revealed no statistically significant difference in comorbidity measures between males and females in the low SR Group (See **Supplemental Table 5 and Supplemental Table 6.**). However, in high SR IBS individuals, females compared to males show greater comorbidity for these comorbidity measures (d=-0.95-1.14), see **Supplemental Table 7 and Supplemental Table 6**.

As an exploratory analyses, potential difference in the comorbidity measures between the Low and High SR HCs were also examined. Even though the means scores on these measures were much lower in HCs as would be expected, HC with high SR compared to Low SR showed moderate effect size increased in the CMSI subscales and the PILL (p’s .02-.09, d=-0.46--0.80; see **Supplemental Tables 8 and 9**)

#### Blood transcriptome profile differences between High and Low SR groups across diagnosis (IBS and HC)

Of the 120 participants providing blood samples, some samples contained insufficient RNA for assay, and some samples also lacked the needed covariate data to yield the covariate-adjusted results. In the final sample of 113 individuals(59 Low SR(23 HC, 40 Female), 54(High SR(12 HC, 36 Female), bioinformatic analyses of all 567 genes showing >1.5-fold differential expression in high-vs. low-SR blood samples (230 up-regulated; 337 down-regulated) linked high SR to increased activity of CREB family of transcription factors involved in mediating beta-adrenergic signaling from the SNS (+0.17 ± SE 0.08 log_2_ TFBM ratio in promoters of genes up-regulated in high-vs. low-SR groups, p = .036) but no difference in the GR transcription factor involved in cortisol signaling from the HPA axis (−0.18 ± 0.25, p = .481). Results also linked high-SR to increased activity of the pro-inflammatory AP-1 family of transcription factors (0.27 ± 0.10, p = .006) but no difference in activity of the pro-inflammatory NF-κB family (−0.08 ± 0.15, p = .605). Among n=46 individual with additional brain imaging data available, the high-SR group also showed significantly elevated average expression of 16 pro-inflammatory gene transcripts (e.g., *IL1B*, IL8/*CXCL8*, COX2/*PTGS2, FOSL2, TNF*; covariate-adjusted difference = 0.25 log_2_ mRNA abundance ± 0.11, t(28) = 2.31, p = 0.029).

#### Whole brain microstructural differences between the High and Low SR groups across diagnosis (Supplemental Tables 10, 11, and 12 and Figures 1 and 2)

Compared to the Low SR group, the combined High SR group (containing both IBS and HC) showed lower MD values in basal ganglia and thalamus, as well as along white matter fiber tracts connecting frontal, temporal and primary sensorimotor cortices. The internal capsule, bilateral cerebral peduncles, the left posterior corona radiata, the left posterior thalamic radiation, the left superior longitudinal fasciculus, and thalamus showed higher FA and lower MD in the High SR group. These findings are consistent with higher fiber or cellular density, increased axon or cell size, and increased preferred directionality of the underlying neuronal structures.

**Figure 1.**
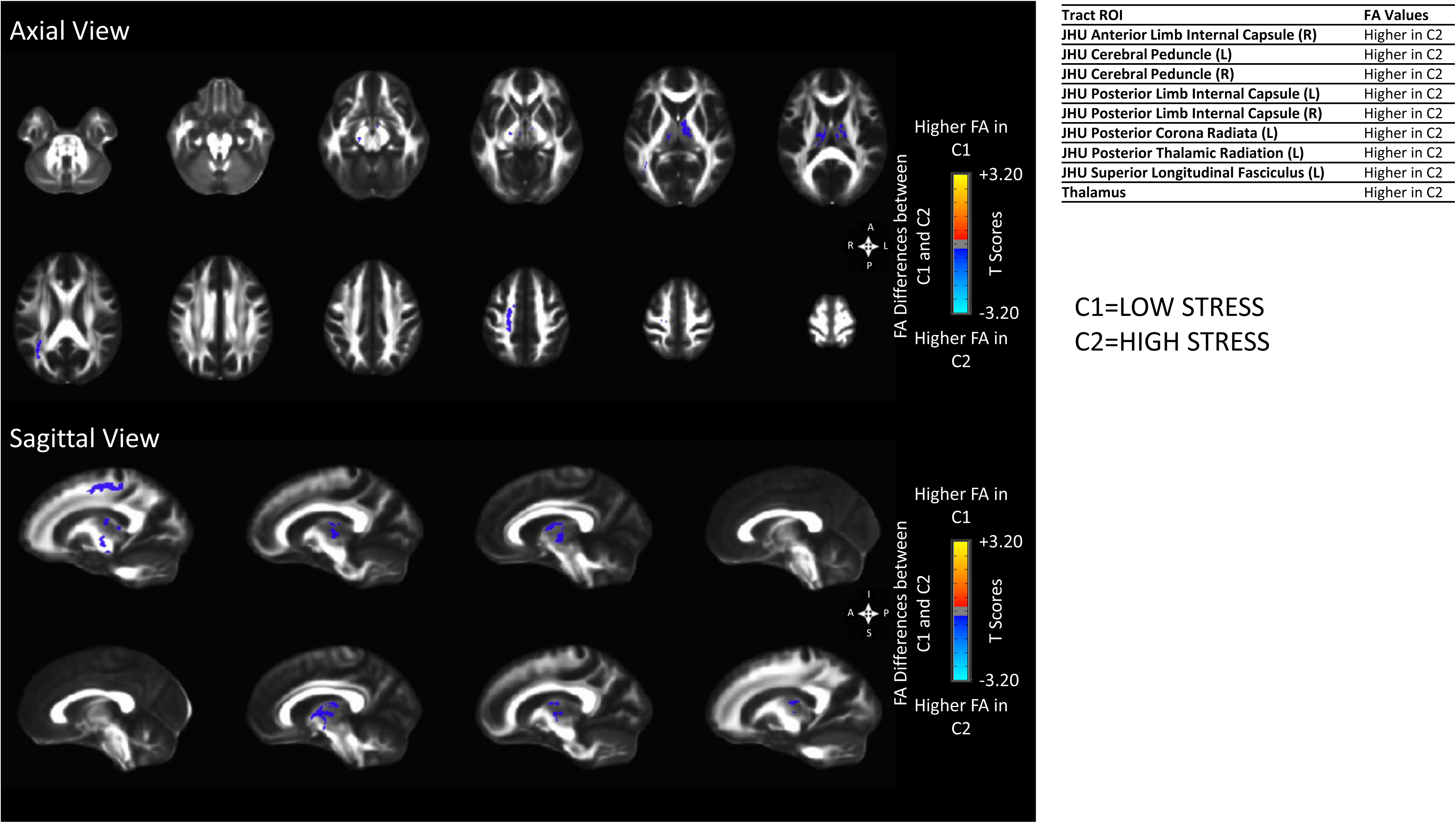
FA Whole Brain Analysis *p*<0.05.

**Figure 2.**
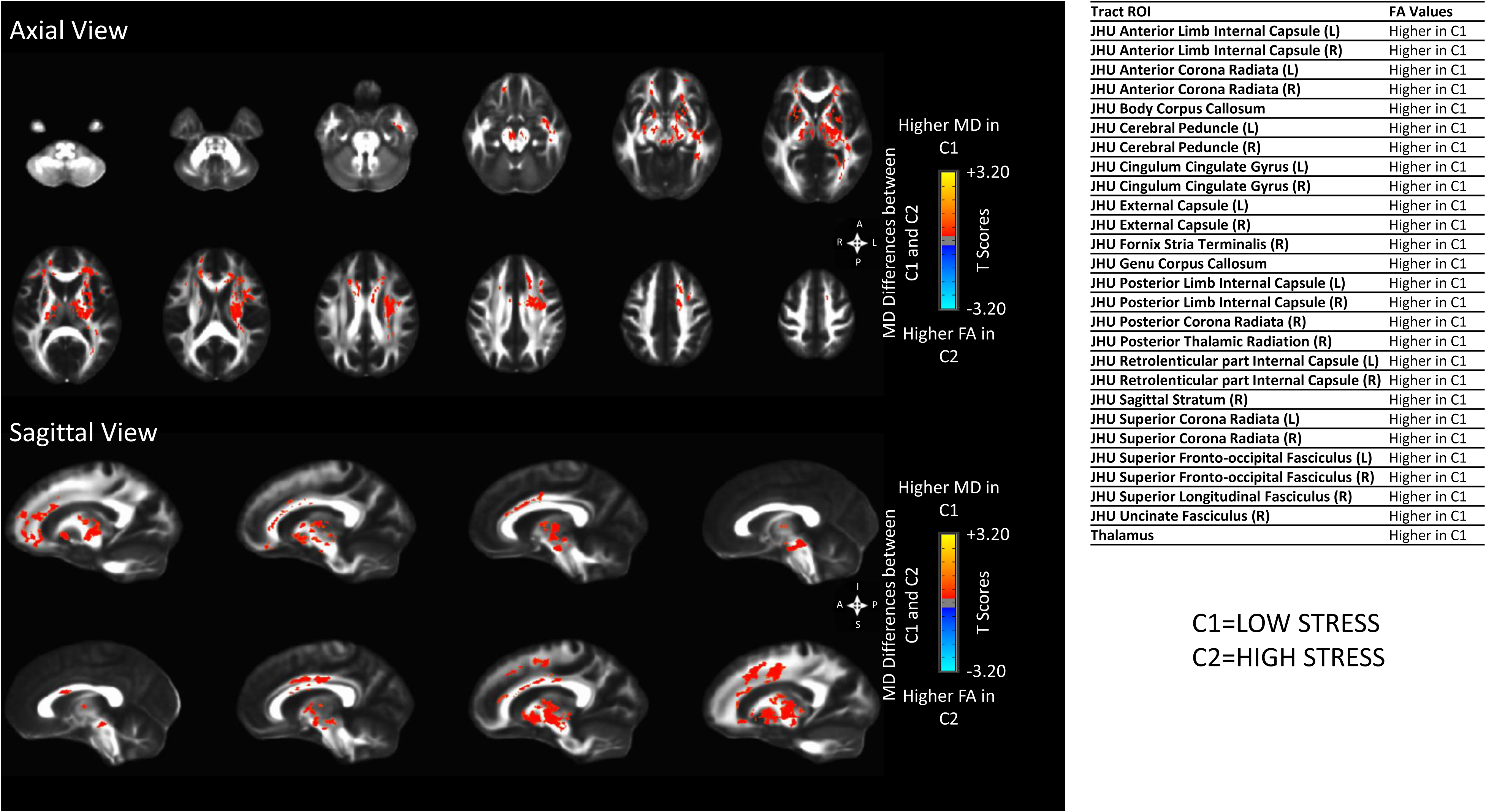
MD Whole Brain Analysis *p*<0.05.

**Brainstem region of interest analysis.** As shown in **Table 2** and **Supplemental Figure 5, 6, 6a**, the High SR group compared to the Low SR group had higher FA and lower MD in the midbrain reticular formation and the ventral tegmental area(VTA). In addition, The MD of the serotonergic dorsal raphe(DRN) and periaqueductal gray (PAG) were also reduced in the High SR group.

**Table 2.** Significant brainstem nuclei showing microstructural difference in low compared to high SR groups.

| Regions | Measure | Direction of the Difference | Cluster Volume ($\mu L$) |
| --- | --- | --- | --- |
| Midbrain Reticular Formation | FA | Higher in High Stress | 43 |
|  | MD | Lower in High Stress | 144 |
| Ventral Tegmental Area | FA | Higher in High Stress | 27 |
|  | MD | Lower in High Stress | 22 |
| Dorsal Raphe | MD | Lower in High Stress | 23 |
| Periaqueductal Gray | MD | Lower in High Stress | 91 |

SR, stress reactivity; FA, fractional anisotropy; $\mu L$ , microliter; MD, mean diffusivity

In summary, based on the microstructural analysis, the High SR group, with a high proportion of more severe IBS participants was characterized by an increase in neuronal or axonal density in sensory processing areas, and brainstem ascending arousal systems as reflected by lower MD and higher FA values.

#### Stress group differences in global connectivity of central autonomic and brainstem regions

Compared to the Low SR, the High SR group showed reductions in the global connectivity of the posterior and anterior insula subregions and regions within the central autonomic network at q<.05 (see Supplemental Table 11). We also observed moderate effect size reductions (Cohen’s d=0.32-0.43) in the High SR group that were significant at nominal p-values (p<.05) in other emotional arousal regions (right hippocampus, right amygdala, bilateral parahippocampal gryus, left rostral anterior cingulate cortex), additional subregions with in the posterior and anterior insula, default mode (bilateral precuneus, left medial prefrontal cortex) regions, as well brain stem nuclei (right parabrachial complex (PBC), median raphe nucleus (MRN).

Furthermore, as shown in **Table 3**, across the groups, reductions in the global connectivity of the posterior and anterior insula were significantly associated with the inflammatory composite score at q<.05 and the CTRA at nominal p values. We also observed reductions of global connectivity in the PBC were associated with increased CTRA at p<.05.

**Table 3.** Association between SR group-specific global resting state connectivity and gene expression composite scores.

| Brain ROI | Description | Region Name | Gene composite Score | B | Std B | SE | t | P | q |
| --- | --- | --- | --- | --- | --- | --- | --- | --- | --- |
| R_InfCirIns | Inferior segment of the circular sulcus of the insula | Posterior Insula | Inflammatory | -2.36 | -0.51 | 0.60 | -3.89 | $3.65 \times 10^{-4}$ | $7.00 \times 10^{-3}$ |
| R_PosLS | Posterior ramus of the lateral sulcus | Posterior Insula | Inflammatory | -2.50 | -0.48 | 0.71 | -3.49 | $1.00 \times 10^{-3}$ | $1.10 \times 10^{-2}$ |
| L_InfCirIns | Inferior segment of the circular sulcus of the insula | Posterior Insula | Inflammatory | -2.14 | -0.42 | 0.70 | -3.03 | $4.00 \times 10^{-3}$ | $2.50 \times 10^{-2}$ |
| R_SupCirIns | Superior segment of the circular sulcus of the insula | anterior Insula | Inflammatory | -1.87 | -0.43 | 0.63 | -2.95 | $5.00 \times 10^{-3}$ | $2.50 \times 10^{-2}$ |
| R_LoInG_CInS | Long insular gyrus and central sulcus of the insula | Posterior Insula | Inflammatory | -1.73 | -0.39 | 0.62 | -2.78 | $8.00 \times 10^{-3}$ | $3.20 \times 10^{-2}$ |
| R_PBC | Parabrachial complex | Parabrachial complex | Inflammatory | -1.11 | -0.37 | 0.43 | -2.61 | $1.30 \times 10^{-2}$ | 0.243 |
| L_InfCirIns | Inferior segment of the circular sulcus of the insula | Posterior Insula | CTRA | -1.80 | -0.34 | 0.83 | -2.18 | $3.50 \times 10^{-2}$ | 0.248 |
| R_InfCirIns | Inferior segment of the circular sulcus of the insula | Posterior Insula | CTRA | -1.60 | -0.33 | 0.75 | -2.13 | $3.90 \times 10^{-2}$ | 0.248 |
| R_SupCirIns | Superior segment of the circular sulcus of the insula | anterior Insula | CTRA | -1.66 | -0.37 | 0.74 | -2.25 | $3.00 \times 10^{-2}$ | 0.248 |

SR, stress reactivity; ROI, region of interest; Std B, standardized beta; SE, standard error; R, right; L, left; CTRA, conserved transcriptional response to adversity.

## Discussion

We were able to identify two subgroups of individuals in a large sample of IBS and HCs which differed in clinical presentation including a history of early life adversity and greater psychosocial comorbidity, microstructural and functional connectivity alterations in the brain and gene expression. The main findings identified a group with higher perception and reactivity to stress (High SR as indexed by scores on IPIP and PSS questionnaires), and greater gene expression for unprovoked SNS activity (as indexed by increased transcription of CREB), and proinflammatory changes in PBMCs. Even though the High SR phenotype was present in both IBS and HCs, it was significantly more common in patients. In IBS participants, SR moderated the sex difference in extraintestinal symptoms. Brain imaging findings showed neuroplastic changes consistent with an upregulation of several brain networks, including ascending arousal systems as well as sensory processing and emotion regulation regions in the brain in addition to functional connectivity changes in the central autonomic network.

### Phenotype of High and Low SR

Our results identified subgroups of IBS and HC participants with High and Low SR with a significantly higher prevalence of the High SR phenotype in IBS than in HC. In both groups, the High SR phenotype was characterized by a history of early life adversity and psychosocial comorbidity, and in IBS with greater symptom severity. We have recently showed that a subgroup of patients with ulcerative colitis who show a High SR phenotype as indexed in the current study, have significantly greater frequency of clinical flares over a 3 year period.^11^ When viewed together with our previous results, one may speculate that a High SR phenotype exists not only in a significant number of patients with disorders of gut brain interaction and with inflammatory bowel disease, where it correlates with disease severity and recurrence of symptoms, but is also present in a smaller number of healthy individuals in which this phenotype may be a risk factor for greater vulnerability to develop IBS and other brain gut disorders, which are more common in women. While there was no significant sex-related difference in the prevalence of high SR, having this phenotype moderated the frequency of extraintestinal symptoms. In other words, non-IBS comorbidities are higher in women with high SR, but not in those without.

### Genomic correlates of High SR

An association between psychological stress and expression of genes bearing response elements for transcription factors that regulate inflammatory and anti-viral processes has previously been reported.^44,45^ In late adolescence, higher levels of daily interpersonal stress and shorter sleep duration were associated with upregulation of genes bearing response elements for pro-inflammatory transcription factor, nuclear factor kappa B (NF-κB).^44^ Furthermore, studies evaluating transcriptome profiles to predict depressive symptoms following an inflammatory challenge with low dose endotoxin suggest that increased baseline activity of transcription factors related to NF-κB and beta-adrenergic signaling (CREB) and decreased activity of GR related transcription factors were associated with post-endotoxin depressed mood.^45^ Using promoter-based bioinformatics analysis, we have previously identified increased activity of CREB transcription factors ^46^ in IBS patients, a population without macroscopic evidence for intestinal inflammation.^47^ The expression of proinflammatory and stress-related genes in individuals without clinical evidence for gut or systemic inflammation suggests the presence of a “genomic memory” encoded by early adverse life events that increases the individual reactivity to psychosocial stress and more severe IBS symptoms.

### Brain signatures correlates of High SR

The High vs Low SR group showed increased density and directionality of microstructure of brain regions involved in sensory processing and integration (basal ganglia and thalamus), and brain stem nuclei involved in ascending arousal systems (midbrain reticular formation, serotonergic dorsal raphe and the VTA). The findings of a decreased MD and increased FA similarly describe an increase in neuronal density and/or structural connectivity into these regions. As the midbrain reticular formation plays a key role in determining which sensory signals reach conscious attention, one may speculate that our findings of higher density/connectivity in this region are a reflection of more pain or sensory information reaching conscious attention in these patients. The clinical implications of the observed microstructural alterations in the VTA are less clear, even though stress-induced increases in VTA excitability have been reported.^48,49^

In addition to the microstructural signatures differentiating the two SR groups, the High SR group showed reductions in the global connectivity of brain regions involved in several brain networks, including the emotional arousal, salience, central autonomic and default mode as well as the dorsal raphe nucleus in the brainstem. Several of these reductions, including insula subregions and parabrachial complex were significantly associated with the inflammatory and CTRA composite scores.

Even though changes in global connectivity in brain regions do not necessarily correlate with the clinical hallmarks of increased SR, e.g. changes in sensory sensitivity and autonomic nervous system responsiveness, the fact that both microstructural and functional connectivity changes were observed in some of the same brain regions, including the serotonergic raphe nucleus is consistent with a significant remodeling of the brain in the high SR phenotype.

### Integrative model of increased stress reactivity

Due to the cross sectional nature of this study, we cannot make any definite conclusions about the causality of factors in the observed association of self-reported stress, brain changes and blood transcription profiles.

However, when viewed together with an extensive preclinical and clinical literature^50,51^, we propose the following model: The High SR phenotype first evolves in the developing brain as a result of interactions between early adversity and genetic vulnerabilities,^52^ resulting in neuroplastic changes in the central autonomic network and an increased tonic activity of the SNS, as well as an increased engagement of both the SNS system and the ascending arousal system to environmental stressors.^53^ The increased SNS activity affects gene expression patterns in bone marrow peripheral blood mononuclear cells (PBMCs), which function like a “genetic memory” of adversity.^54^ During acute stressors, even if mild, primed PBMCs can enter the systemic circulation and reach the brain, where they induce neuroplastic changes in brainstem nuclei and several brain networks involved the sensory processing and integration regions, contributing to the visceral hypersensitivity observed in a significant subset of patients^55,56^. Evidence for an association of peripheral immune cell priming and the presence of widespread pain and comorbidities has been reported in patients with chronic pelvic pain, a condition often comorbid with IBS.^57^ Alternatively, the neuroplastic changes in the sensory and emotion regulation networks of the brain could result from the interactions of vulnerability genes and early life adversities even without the stress-induced inflammatory feedback from the bone marrow.

### Clinical Implications

We demonstrate that a High SR clinical and biological phenotype in both IBS patients and HCs can be identified simply by using specific cutoffs on two well established questionnaires, the PSS and the IPIP-N. This phenotyping may have significant clinical implications. In patients presenting with new, unexplained symptoms of abdominal pain and altered bowel habits following a stressful life period or traumatic event, or following an enteric infection phenotyping may help to identify individuals at higher risk for developing IBS. In IBS, the presence of a high SR phenotype may help to design personalized treatment approaches. Those with High SR may show greater benefit from beta blockers.^58^ Beta-adrenergic blockade has been shown to blunt inflammatory and antiviral/ antibody gene expression responses to acute psychosocial stress in animal models. Even though therapeutic outcomes from such treatments in unselected IBS patients have been mixed, beta blockers might be considered in High SR patients where stress and autonomic dysfunction are prominent features contributing to symptoms. The High SR subgroup may show greater benefits from brain gut behavior therapies, such as mindfulness-based stress reduction, gut directed cognitive behavioral therapy (CBT) or hypnotherapy aimed to mitigate enhanced stress responsiveness even though subgroup analyses based on SR should be performed all treatment trials. Behavioral treatments have been shown to reliably reduce perceptions of stress in both HCs^59,60^ and IBS groups with IBS patients showing greater benefits such as durable decreases GI and extraintestinal symptoms^61^ and enhanced QOL,^62^ but it remains to be determined if such treatments also normalize the increased SNS tone. Importantly, early interventions, including parental training has been shown to prevent /reduce stress-related psychopathology in at-risk children^63,64^ and could feasibly delay or prevent neuroplastic brain changes and the development of the High SR phenotype or IBS, although further studies are needed.

## Supporting information

Supplemental methods and will be used for the link to the file on the preprint site.

Supplemental methods and will be used for the link to the file on the preprint site.

## Data Availability

The 16S sequences and associated metadata used in this study are part of several ongoing projects within our research group. Due to the active nature of these projects and our ongoing efforts in writing and publishing additional papers, we are not able to publicly release the entire dataset at this time. However, we are committed to transparency and scientific collaboration. Therefore, specific data requests can be accommodated on a case-by-case basis. We plan to submit the sequence and associated metadata to an appropriate publicly available archive once all ongoing projects are completed.

## Acknowledgements

We acknowledge the services of the U54 SCORE Data Processing and Analysis Core, the Neuroimaging and the Bioinformatics Cores of the Goodman Luskin Microbiome Center. Special thanks to Dr. Annie Gupta for providing funds to perform the sequencing of the PBMC samples. We thank Cathy Liu and Madelaine Leitman for valuable editorial assistance.

---

**Supplemental Figure 1.**
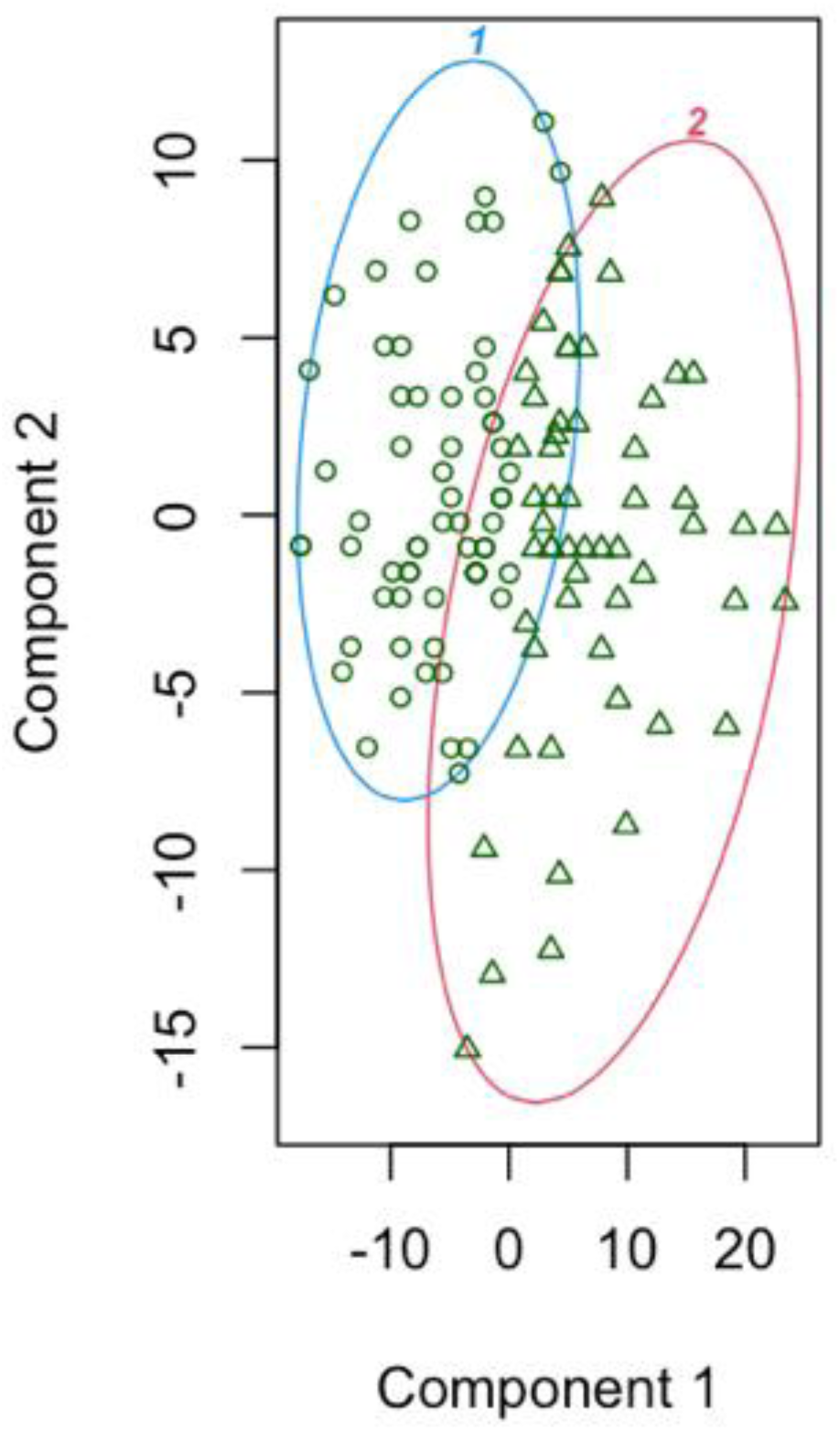
Clustering participants with blood samples in stress reactivity groups.

**Supplemental Figure 2.**
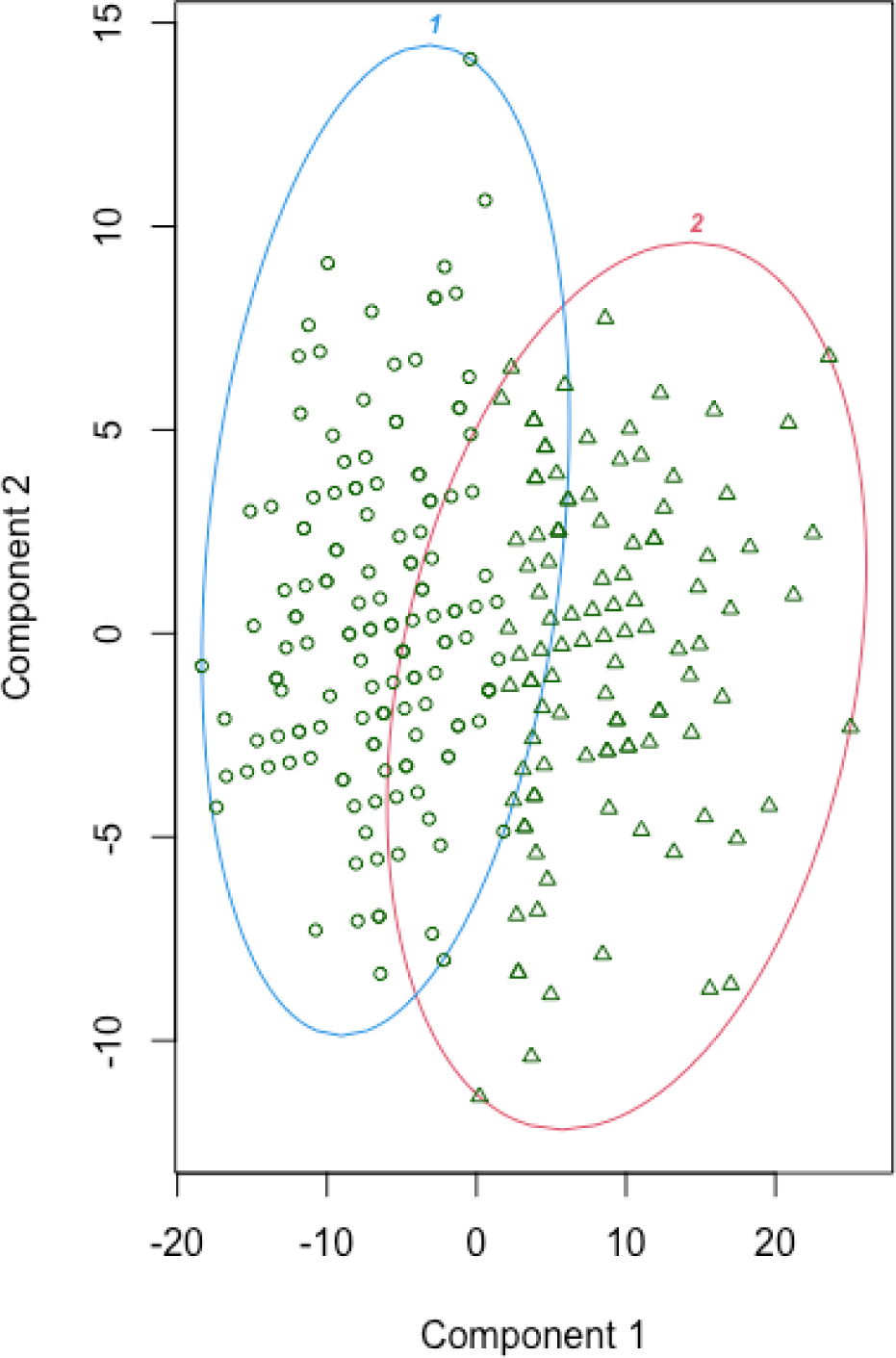
Clustering Participants with Brain Imaging Data in Stress Reactivity Groups.

**Supplemental Figure 3.**
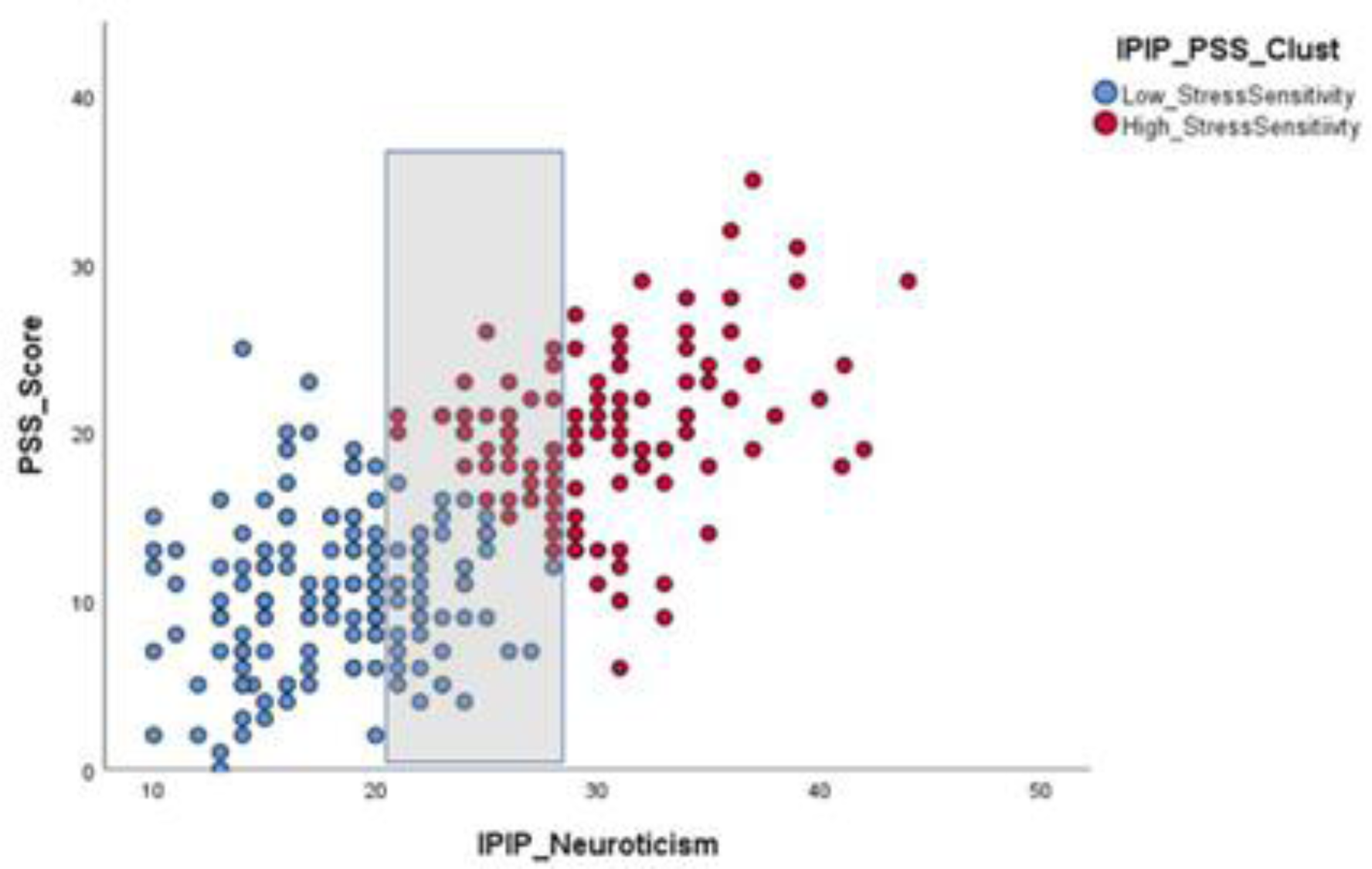
Overlap in IPIP-N in Low and High SR Groups.

**Supplemental Figure 4.**
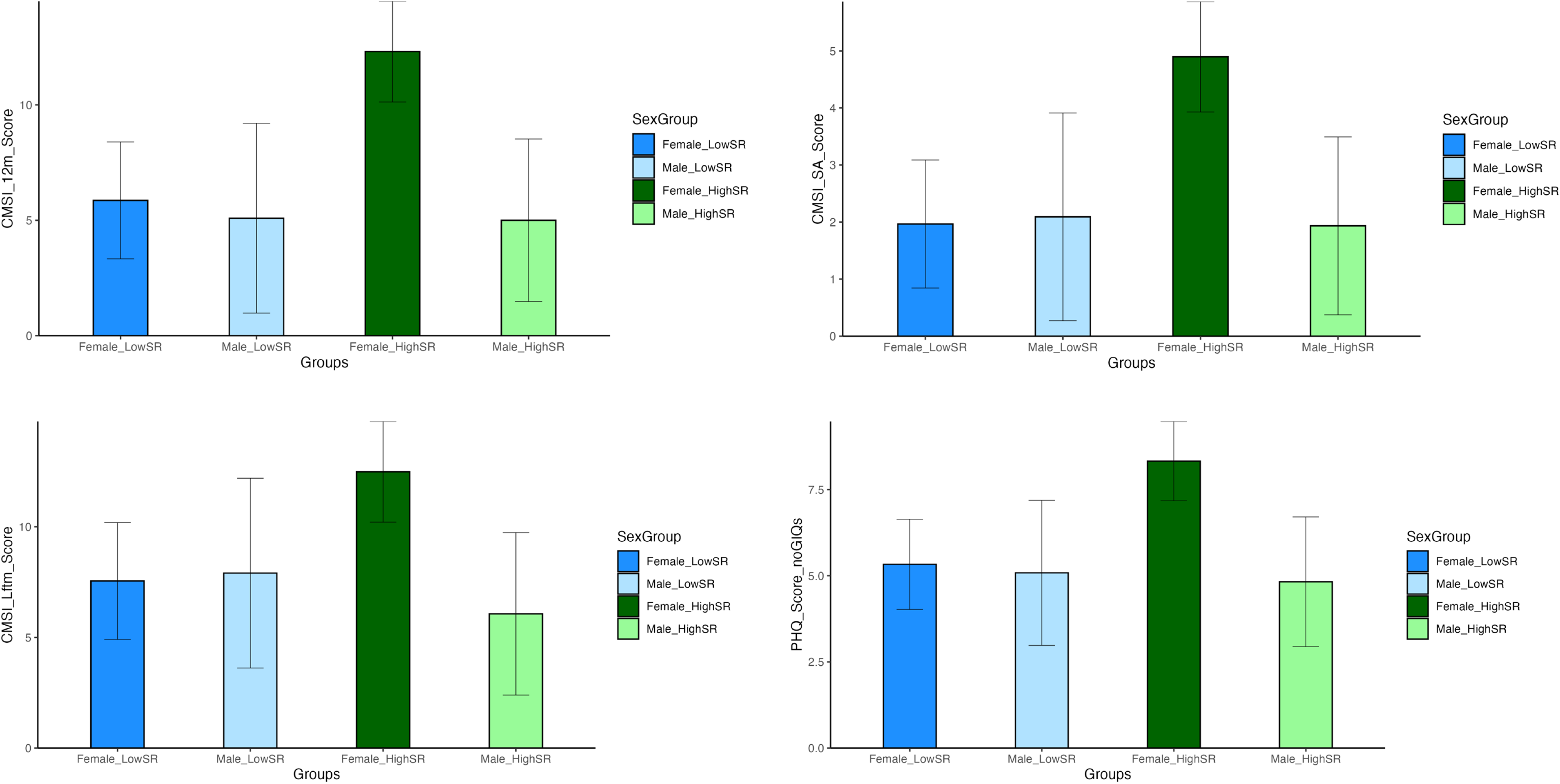
Box plots showing comorbidity scores by sex and SR group.

**Supplemental Figure 5.**
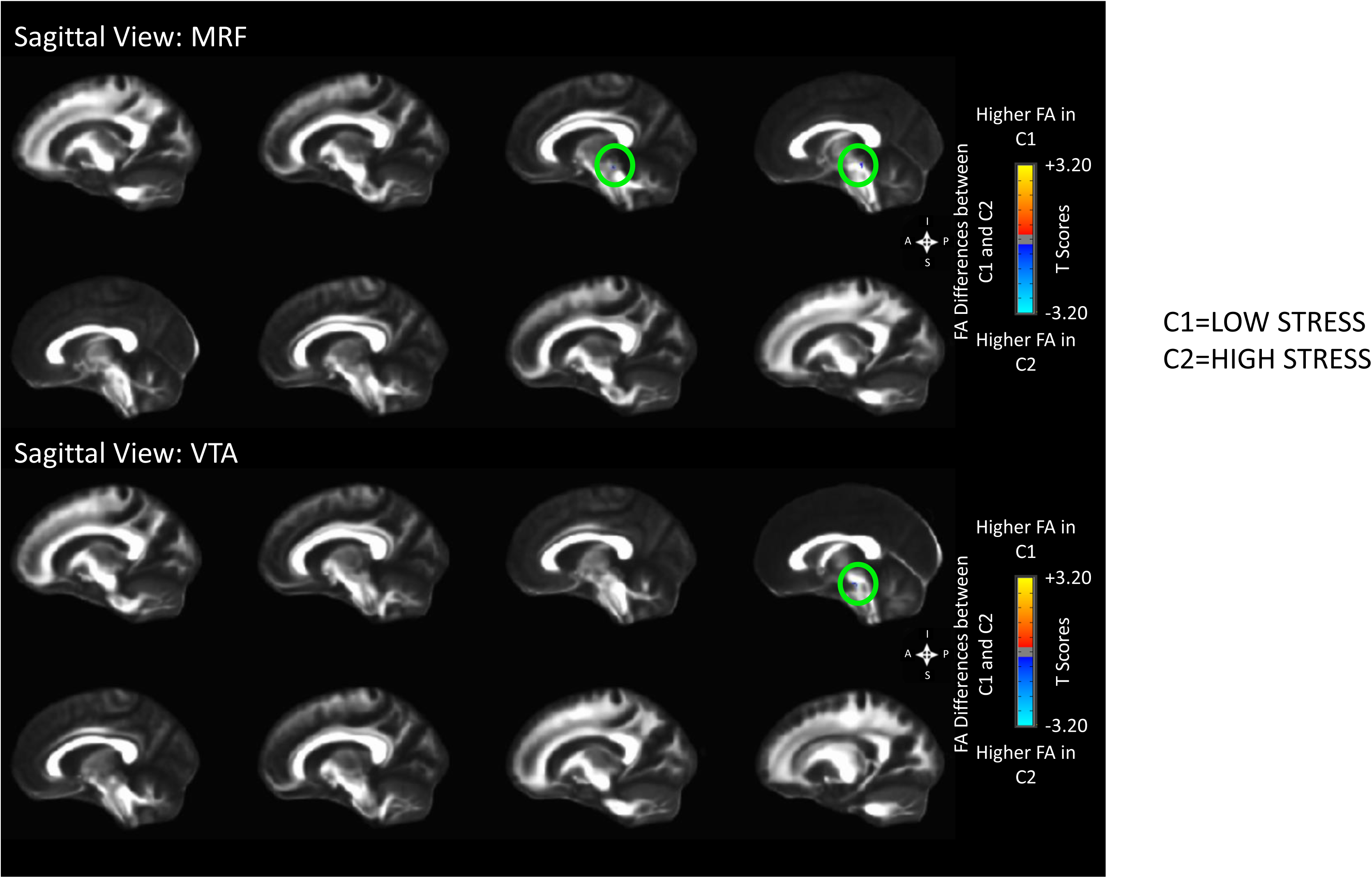
FA ROI Analysis *p*<0.05.

**Supplemental Figure 6.**
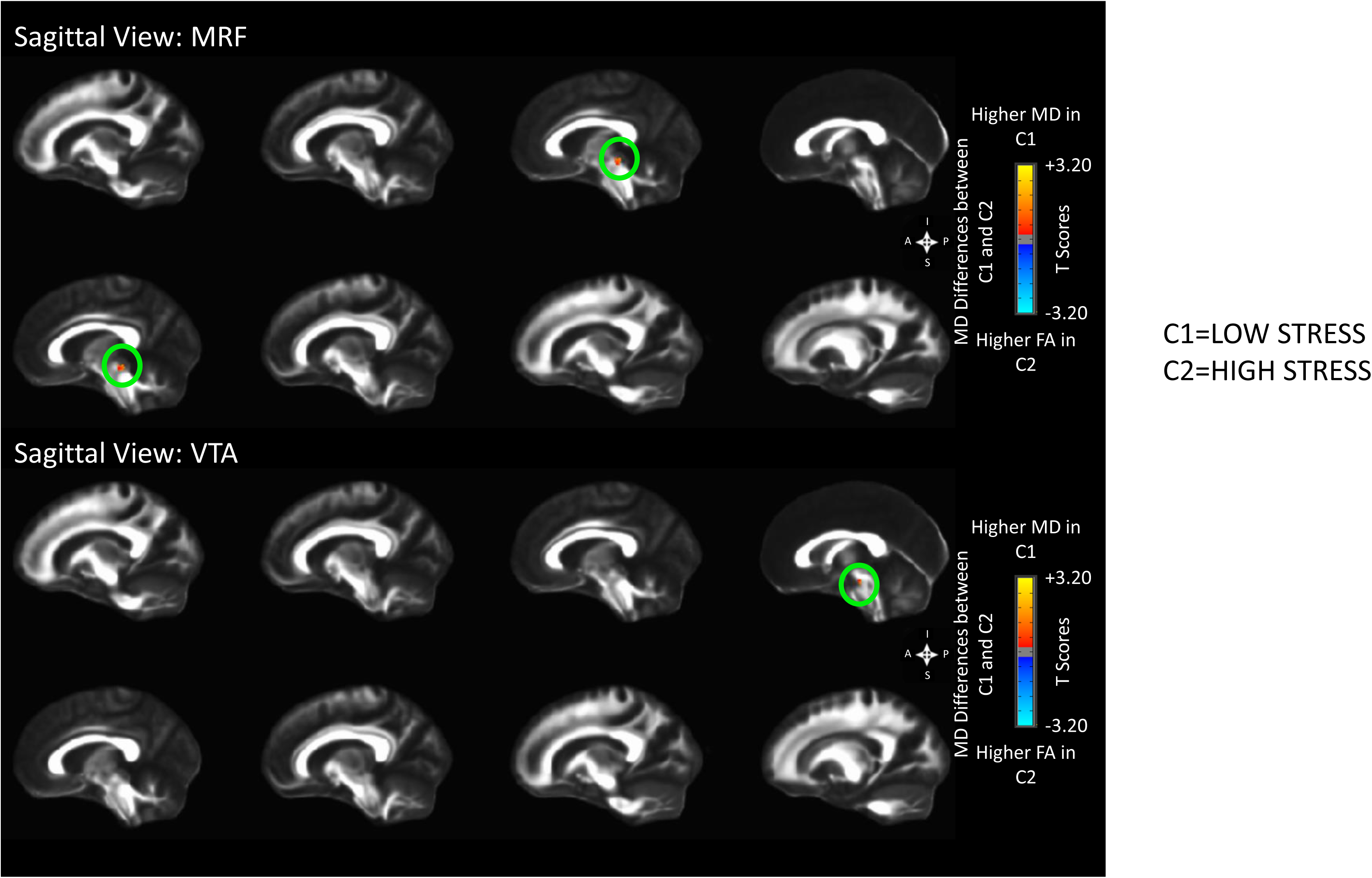
MD ROI Analysis *p*<0.05.

**Supplemental Figure 6a.**
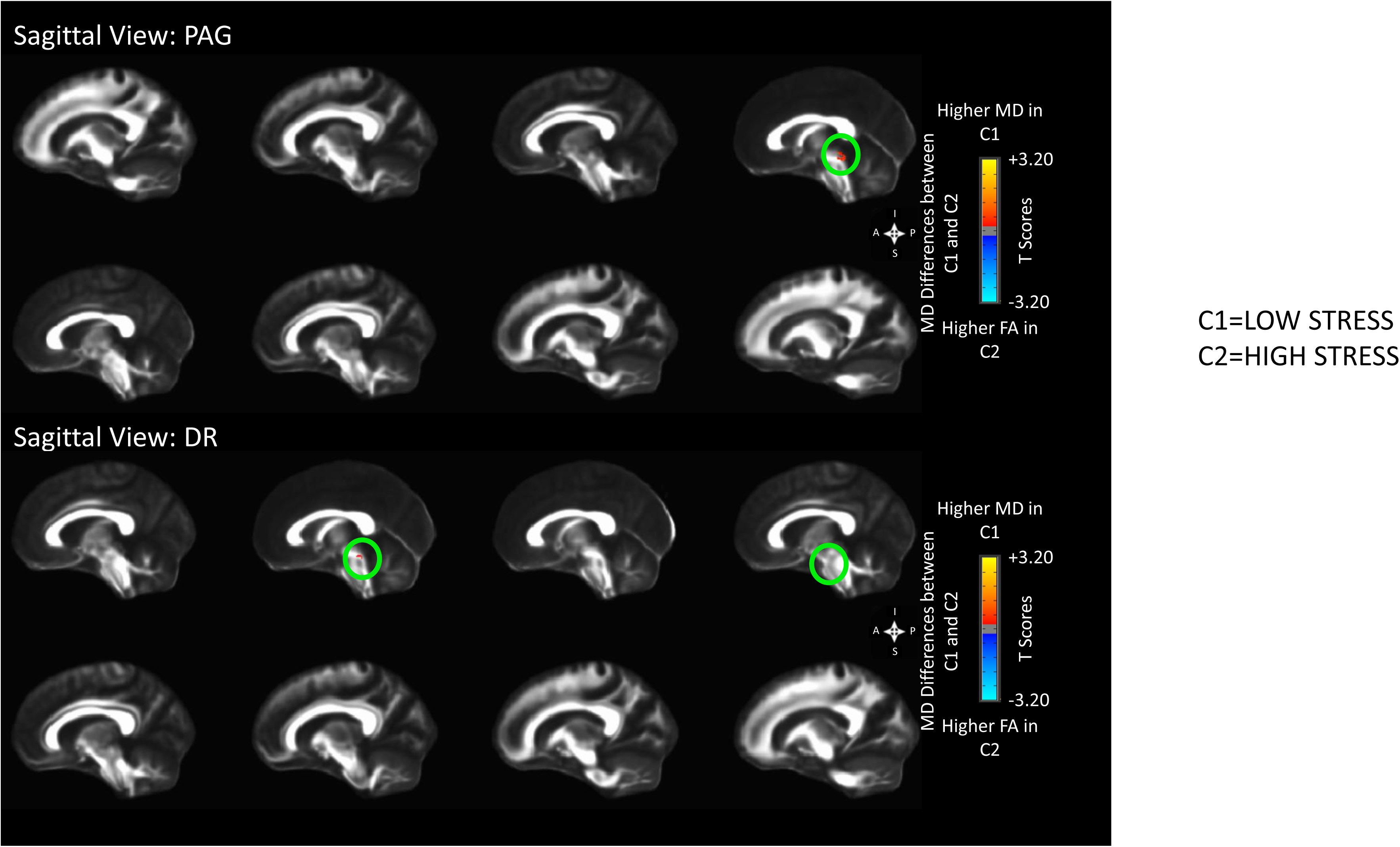
MD ROI Analysis *p*<0.05.

