## Supplemental methods and will be used for the link to the file on the preprint site. for "IBS stress reactivity phenotype is associated with blood transcriptome profiles and microstructural and functional brain changes"

**Blood RNA profiling.**  Total RNA was isolated from whole blood samples (PAXgene RNA tubes), tested for suitable mass, converted to cDNA using a high-efficiency mRNA-targeted enzyme system (Lexogen QuantSeq 3’ FWD), and sequenced on an Illumina HiSeq 4000 instrument in the UCLA Neuroscience Genomics Core Laboratory. Assays targeted > 10 million sequencing reads per sample (achieved median=16 million), each of which was mapped to the GRCh38 reference human transcriptome using the STAR aligner (median mapping rate=93.4%) and quantified as gene-specific reads per million mapped reads.

**Diffusion Tensor Imaging (DTI).** DTIs were collected in 64 equidistant diffusion-sensitizing directions with b=1000s/mm^2^, along with a single b=0s/mm^2^ image, with echo time (TE)/repetition time (TR)=88ms/9500ms; matrix size=128×128; field of view (FOV)=256mm; and a slice thickness=2mm, with no interslice gap, resulting in 2mm isotropic resolution. All subject DTI data was resampled to 1 mm isotropic resolution for analyses in standard space. Fractional Anisotropy (FA) and mean diffusivity (MD) images for each participant were registered to the Johns Hopkins University DTI atlas (ICBM-DTI-81 1mm FA atlas) using the FLIRT and FNIRT commands in FSL.

*DTI Processing and Computational Statistics.* DTI data were processed using methodology similar to previous studies.^14,34-36^ Briefly, diffusion MRI scans were corrected for eddy currents and motion using the eddy correct functionality of the FSL Diffusion Toolbox (FDT) as part of FSL (FMRIB; Oxford, UK).^37^ Following skull extraction using the Brain Extraction Tool (BET), fractional Anisotropy (FA) and mean diffusivity (MD) values at the voxel level were calculated for DTI, within a mask consisting of both cortical white matter and deep gray matter regions of interest, including the basal ganglia and thalamus, (namely the basal ganglia and thalamus, defined by their respective masks from the Harvard-Oxford subcortical atlas) as defined previously.^14,34^ All FA and MD images for each participant were registered to the Johns Hopkins University DTI atlas (ICBM-DTI-81 1mm FA atlas) using the linear (12 directions via FLIRT) and non-linear (FNIRT) commands in FSL.

**Resting state imaging.** Resting-state scans (eyes closed and noise-reducing headphones) were acquired with the following parameters: 40-slice whole brain volumes, slice thickness = 4mm, repetition time = 2000ms, echo time= 28ms, resting acquisition time = 10m6s, flip angle = 77°, field of view = 220, 2×2×2 mm voxel size. A high-resolution structural image was obtained from each subject for registration purposes with a magnetization-prepared rapid acquisition gradient-echo sequence, repetition time = 2200ms, echo time = 3.26ms, structural acquisition time =5m 12s, slice thickness = 1mm, 176 slices, 256*256 voxel matrix, 1mm voxel size.

**Resting state processing and computational analyses**. Using established computational pipelines,^39-41^ we applied graph theory to the resting state data to compute the connectivity strength or global influence of brain regions comprising the central autonomic network (CAN) and brainstem regions of interest on brain functioning. Strength represents the weighted sum of connections (i.e., Fisher Z transformed correlations) for each region of interest. High compared to low values for strength indicate greater influence on the global state of brain functioning. As defined by the Destrieux (cortical)^42^ and Harvard-Oxford Subcortical Atlases^43^, and the Harvard Ascending Arousal Network (AAN) Atlas, a priori specified individual regions of interested included the posterior insula (long insular gyrus and central sulcus of the insula, and the inferior segment of the circular sulcus of the insula, posterior ramus of the lateral sulcus), anterior insula (short insular gyri, anterior segment of the circular sulcus of the insula, Superior segment of the circular sulcus of the insula, horizontal ramus of the anterior segment of the lateral sulcus, and the vertical ramus of the anterior segment of the lateral sulcus), medial prefrontal cortex (defined as the mean of the orbital gyrus, orbital sulcus, lateral orbital sulcus, medial orbital sulcus, suborbital gyrus, rectus gyrus and the transverse frontopolar gyri and sulci), precuneus, Anterior part of the cingulate gyrus and sulcus, middle-anterior part of the cingulate gyrus and sulcus, thalamus, amygdala, para hippocampal gyrus, hippocampus, cerebellum, PAG, PBC, and VTA. We also included the LC and DRN and MRN.
