## Supplemental methods and will be used for the link to the file on the preprint site. for "IBS stress reactivity phenotype is associated with blood transcriptome profiles and microstructural and functional brain changes"

Supplemental Table 1. Means Within and Across Diagnosis on Demographic and Psychosocial Variables

| Variables | Across Diagnosis | | HC | | IBS | |
|---|---|---|---|---|---|---|
| | Low SR *n* = 100 *M(SD)* | High SR *n* = 68 *M(SD)* | Low SR *n* = 57 *M(SD)* | High SR *n* = 13 *M(SD)* | Low SR *n* = 43 *M(SD)* | High SR *n* = 55 *M(SD)* |
| ACE Total Score | 1.09(1.30) | 2.01(1.78) | 0.88(1.32) | 2.23(2.20) | 1.37(1.24) | 1.96(1.68) |
| Age | 30.61(11.84) | 28.54(10.27) | 31.33(11.34) | 28.77(13.72) | 29.65(12.53) | 28.49(9.44) |
| BMI | 24.60(3.35) | 23.51(3.17) | 25.42(3.41) | 26.17(3.69) | 23.52(2.96) | 22.88(2.71) |
| BRS Score | 24.29(3.51) | 17.93(4.93) | 24.61(3.19) | 19.75(5.31) | 23.86(3.88) | 17.53(4.80) |
| CDRISC Adaptability/Ability to Bounce Back | 17.85(2.09) | 13.88(3.24) | 17.89(1.89) | 14.23(3.27) | 17.79(2.37) | 13.80(3.26) |
| CDRISC Emotional/Cognitive Control Under Pressure | 21.83(3.45) | 17.26(4.48) | 21.85(3.35) | 17.00(4.74) | 21.81(3.61) | 17.33(4.46) |
| CDRISC Meaning/Spiritual | 5.45(2.29) | 3.56(2.57) | 5.43(2.36) | 5.15(2.19) | 5.48(2.22) | 3.18(2.52) |
| CDRISC Persistence/Tenacity and Self-efficacy | 27.82(3.29) | 21.02(5.45) | 27.60(3.21) | 19.46(4.31) | 28.12(3.42) | 21.38(5.66) |
| CDRISC Meaning/Control | 10.26(1.52) | 7.06(2.38) | 10.04(1.65) | 7.62(2.47) | 10.56(1.30) | 6.93(2.36) |
| CSQ Catastrophizing Score | 0.67(0.95) | 1.68(1.32) | 0.22(0.40) | 0.88(0.84) | 1.27(1.26) | 1.86(1.35) |
| Early Trauma Inventory - Total Score | 3.34(3.39) | 5.94(4.53) | 2.74(3.03) | 5.54(4.33) | 4.17(3.68) | 6.04(4.61) |
| HAD Anxiety | 3.26(3.15) | 9.18(4.26) | 2.04(2.35) | 7.62(3.33) | 4.87(3.37) | 9.55(4.40) |
| IPIP Neuroticism | 16.19(2.93) | 33.05(3.62) | 15.81(2.98) | 32.62(2.66) | 16.71(2.82) | 33.15(3.82) |
| PSS Score | 10.50(4.97) | 20.27(5.92) | 9.44(5.00) | 18.15(7.21) | 11.91(4.62) | 20.78(5.53) |
| STAI-T Anxiety | 43.03(6.07) | 62.15(10.74) | 42.26(5.82) | 60.00(11.07) | 44.05(6.59) | 62.65(10.70) |
| VSI Score | 18.29(20.80) | 35.72(20.70) | 3.04(4.29) | 6.77(7.29) | 38.51(16.16) | 42.57(16.43) |

SR, stress reactivity; IBS, irritable bowel syndrome; HC, healthy control; *M*, mean; *SD*, standard deviation; ACE, Adverse Childhood Experience survey; BMI, body mass index; BRS, Brief Resilience Scale; CDRISC, Conner-Davidson Resilience Scale; CSQ, Coping Strategies Questionnaire; HAD, Hospital Anxiety and Depression Scale; IPIP, International Personality Item Pool; PSS, perceived stress scale; STAI-T, Spielberger State Trait Anxiety Inventory-Trait; VSI, Visceral Sensitivity Index.

Supplemental Table 2. Within IBS and HC Group Comparison of Low Versus High SR Groups on Demographic and Psychosocial Variables

| Variables | HC | | | | IBS | | | |
|---|---|---|---|---|---|---|---|---|
| | t | df | *P* | Cohen's d (95% CIs) | t | df | *P* | Cohen's d (95% CIs) |
| ACE Total Score | -2.90 | 68 | 0.01 | -0.89 (-1.51- -0.27) | -1.93 | 96 | 0.06 | -0.39 (-0.80-0.01) |
| Age | 0.71 | 68 | 0.48 | 0.22 (-0.39-0.82) | 0.52 | 96 | 0.60 | 0.11 (-0.29-0.51) |
| BMI | -0.71 | 68 | 0.48 | -0.22 (-0.82-0.37) | 1.11 | 96 | 0.27 | 0.23 (-0.18-.63) |
| BRS Score | 4.23 | 67 | $7.35 \times 10^{-5}$ | 1.34 (0.68-2.00) | 7.04 | 96 | $2.92 \times 10^{-10}$ | 1.43 (0.98-1.88) |
| CDRISC Adaptability/Ability to Bounce Back | 5.46 | 68 | $7.15 \times 10^{-7}$ | 1.68 (1.01-2.34) | 6.75 | 96 | $1.13 \times 10^{-9}$ | 1.37 (0.93-1.82) |
| CDRISC Emotional/Cognitive Control Under Pressure | 4.34 | 68 | $4.82 \times 10^{-5}$ | 1.33 (0.69-1.97) | 5.36 | 96 | $5.70 \times 10^{-7}$ | 1.09 (0.66-1.52) |
| CDRISC Meaning/Spiritual | 0.38 | 67 | 0.70 | 0.12 (-0.49-0.72) | 4.68 | 95 | $9.66 \times 10^{-6}$ | 0.96 (0.53-1.38) |
| CDRISC Persistence/Tenacity and Self-efficacy | 7.72 | 68 | $7.02 \times 10^{-11}$ | 2.37 (1.65-3.09) | 6.88 | 96 | $6.21 \times 10^{-10}$ | 1.40 (0.95-1.84) |
| CDRISC Meaning/Control | 4.33 | 68 | $5.06 \times 10^{-5}$ | 1.33 (0.68-1.97) | 9.05 | 95 | $1.77 \times 10^{-14}$ | 1.85 (1.37-2.33) |
| CSQ Catastrophizing Score | -4.13 | 65 | $1.07 \times 10^{-4}$ | -1.32 (-1.98- -0.65) | -2.31 | 94 | 0.02 | -0.47(-0.88- -0.06) |
| Early Trauma Inventory - Total Score | -2.76 | 68 | 0.01 | -0.85 (-1.47- -0.23) | -2.15 | 94 | 0.03 | -0.44 (-0.85- -0.03) |
| HAD Anxiety | -7.11 | 68 | $8.87 \times 10^{-10}$ | -2.19 (-2.89- -1.48) | -5.77 | 96 | $9.79 \times 10^{-8}$ | -1.17 (-1.60- -0.74) |
| IPIP Neuroticism | -18.66 | 68 | $1.86 \times 10^{-28}$ | -5.74 (-6.86—4.59) | -23.63 | 96 | $8.61 \times 10^{-42}$ | -4.81 (-5.59- -4.02) |
| PSS Score | -5.20 | 68 | $2.03 \times 10^{-6}$ | -1.60 (-2.25- -0.93) | -8.46 | 96 | $3.03 \times 10^{-13}$ | -1.72 (-2.19- -1.25) |
| STAI-T Anxiety | -8.39 | 68 | $4.25 \times 10^{-12}$ | -2.58 (-3.32- -1.83) | -10.01 | 96 | $1.42 \times 10^{-16}$ | -2.04 (-2.53- -1.54) |
| VSI Score | -2.45 | 68 | 0.02 | -0.75 (-1.37- -0.14) | -1.22 | 96 | 0.23 | -0.25 (-0.65-0.15) |

SR, stress reactivity; IBS, irritable bowel syndrome; HC, healthy control; ACE, Adverse Childhood Experience survey; BMI, body mass index; BRS, Brief Resilience Scale; CDRISC, Conner-Davidson Resilience Scale; CSQ, Coping Strategies Questionnaire; HAD, Hospital Anxiety and Depression Scale; IPIP, International Personality Item Pool; PSS, perceived stress scale; STAI-T, Spielberger State Trait Anxiety Inventory-Trait; VSI, Visceral Sensitivity Index.

Supplemental Table 3. Differences in Symptom and Comorbidity Measures between IBS in the Low Compared to the High SR Group

| Variable | t | df | *P* | Cohen's d (95%CIs) |
|---|---|---|---|---|
| IBS-SS Score [past 10 days] | -2.60 | 92 | 0.011 | -.54(-.95--0.12) |
| IBSQoL Total Score [past month] | 2.39 | 93 | 0.011 | 0.49(.08-.90) |
| PILL Score | -4.41 | 96 | $2.7 \times 10^{-5}$ | -0.9 (-1.32--0.47) |
| CMSI_SA_Score | -3.13 | 92 | $2.3 \times 10^{-3}$ | -0.65 (-1.08--0.23) |
| CMSI_12m_Score | -3.06 | 92 | $2.9 \times 10^{-3}$ | -0.64 (-1.06--0.21) |
| PHQ_Score | -2.71 | 96 | $7.9 \times 10^{-3}$ | -0.55 (-0.96--0.14) |
| CMSI_Lft_Score | -1.97 | 92 | 0.05 | -0.41 (-0.83-0.01) |
| CMSI_SS_Score | -1.86 | 92 | 0.07 | -0.39 (-0.81-0.03) |

IBS, irritable bowel syndrome; SR, stress reactivity; IBS-SS, Irritable Bowel Symptom Severity; IBSQoL, Irritable Bowel Syndrome Quality of Life Instrument; PILL, Pennebaker Inventory of Limbic Languidness; CMSI_SA_Score, Complex Medical Symptom Inventory Somatic Awareness; CMSI_12m_Score, Complex Medical Symptom Inventory reporting presence of somatic symptoms for 3 months out of the past year; PHQ_Score, Patient Health Questionnaire without gastrointestinal items; CMSI_Lftm_Score, Complex Medical Symptom Inventory reporting presence of somatic symptoms over the lifetime; CMSI_SS_Score, Complex Medical Symptom Inventory Sensory Sensitivity.

Supplemental Table 4. Means by High and Low SR Groups in IBS

| Variable | Low SR | | | High SR | | |
|---|---|---|---|---|---|---|
| | M | SE | n | M | SE | n |
| IBS-SS Score [past 10 days] | 206.07 | 12.77 | 42 | 251.98 | 12.10 | 52 |
| IBSQoL Total Score [past month] | 68.66 | 2.86 | 42 | 58.38 | 3.08 | 53 |
| PILL Score | 13.40 | 1.09 | 43 | 20.31 | 1.09 | 55 |
| CMSI_SA_Score | 2.00 | 0.29 | 40 | 4.07 | 0.53 | 54 |
| CMSI_12m_Score | 5.65 | 0.79 | 40 | 4.07 | 1.16 | 54 |
| PHQ_Score | 5.26 | 0.59 | 43 | 7.37 | 0.51 | 55 |
| CMSI_Lft_Score | 7.65 | 0.99 | 40 | 10.70 | 1.11 | 54 |
| CMSI_SS_Score | 0.43 | 0.14 | 40 | 0.80 | 0.14 | 54 |

IBS, irritable bowel syndrome; SR, stress reactivity; IBS-SS, Irritable Bowel Symptom Severity; IBSQoL, Irritable Bowel Syndrome Quality of Life Instrument; PILL, Pennebaker Inventory of Limbic Languidness; CMSI_SA_Score, Complex Medical Symptom Inventory Somatic Awareness; CMSI_12m_Score, Complex Medical Symptom Inventory reporting presence of somatic symptoms for 3 months out of the past year; PHQ_Score, Patient Health Questionnaire without gastrointestinal items; CMSI_Lftm_Score, Complex Medical Symptom Inventory reporting presence of somatic symptoms over the lifetime; CMSI_SS_Score, Complex Medical Symptom Inventory Sensory Sensitivity.

Supplemental Table 5. Male - Female Comorbidity Differences in the Low SR Group

| Variables | estimate | SE | df | t | *P* | q | Cohen's d (95% CIs) |
|---|---|---|---|---|---|---|---|
| CMSI_SS_Score | -0.59 | 0.31 | 90 | -1.87 | 0.07 | 0.39 | -0.66 (-1.37-0.05) |
| PHQ_Score | -0.25 | 1.25 | 94 | -0.20 | 0.84 | 0.91 | -0.07 (-0.74- 0.61) |
| PILL_Score | -2.51 | 2.57 | 94 | -0.98 | 0.33 | 0.91 | -0.33 (-1.01-0.34) |
| CMSI_12m_Score | -0.77 | 2.43 | 90 | -0.32 | 0.75 | 0.91 | -0.11 (-0.82-0.59) |
| CMSI_Lftm_Score | 0.36 | 2.53 | 90 | 0.14 | 0.89 | 0.91 | 0.05 (-0.65-0.75) |
| CMSI_SA_Score | 0.13 | 1.08 | 90 | 0.12 | 0.91 | 0.91 | 0.04 (-0.66-0.74) |

SR, Stress Reactivity; SE, standard error; df, degrees of freedom; CMSI_SS_Score, Complex Medical Symptom Inventory Sensory Sensitivity; PHQ_Score, Patient Health Questionnaire without gastrointestinal items; PILL, Pennebaker Inventory of Limbic Languidness; CMSI_12m_Score, Complex Medical Symptom Inventory reporting presence of somatic symptoms for 3 months out of the past year; CMSI_Lftm_Score, Complex Medical Symptom Inventory reporting presence of somatic symptoms over the lifetime; CMSI_SA_Score, Complex Medical Symptom Inventory Somatic Awareness

Supplemental Table 6. Estimated Means by Sex and SR Group

| Variables | Male Low SR | | Female Low SR | | Male High SR | | Female High SR | |
|---|---|---|---|---|---|---|---|---|
| | *M* | SE | *M* | SE | *M* | SE | *M* | SE |
| CMSI_SS_Score | -8E-16 | 0.27 | 0.59 | 0.16 | 0.07 | 0.23 | 1.08 | 0.14 |
| PHQ_Score | 5.08 | 1.06 | 5.33 | 0.66 | 4.82 | 0.95 | 8.33 | 0.58 |
| PILL_Score | 11.58 | 2.18 | 14.10 | 1.36 | 16.47 | 1.95 | 21.76 | 1.19 |
| CMSI_12m_Score | 5.09 | 2.07 | 5.86 | 1.27 | 5.00 | 1.77 | 12.31 | 1.10 |
| CMSI_Lftm_Score | 7.91 | 2.16 | 7.55 | 1.33 | 6.07 | 1.85 | 12.49 | 1.15 |
| CMSI_SA_Score | 2.09 | 0.92 | 1.97 | 0.56 | 1.93 | 0.79 | 4.90 | 0.49 |

SR, Stress Reactivity; M, means; SE, standard error; CMSI_SS_Score, Complex Medical Symptom Inventory Sensory Sensitivity; PHQ_Score, Patient Health Questionnaire without gastrointestinal items; PILL, Pennebaker Inventory of Limbic Languidness; CMSI_12m_Score, Complex Medical Symptom Inventory reporting presence of somatic symptoms for 3 months out of the past year; CMSI_Lftm_Score, Complex Medical Symptom Inventory reporting presence of somatic symptoms over the lifetime; CMSI_SA_Score, Complex Medical Symptom Inventory Somatic Awareness

Supplemental Table 7. Male - Female Comorbidity Differences in the High SR Group

| Variables | estimate | SE | t | df | *P* | q | Cohen's d (95% CIs) |
|---|---|---|---|---|---|---|---|
| CMSI_SS_Score | -1.01 | 0.27 | -3.75 | 90 | $3 \times 10^{-4}$ | $1.9 \times 10^{-3}$ | -1.14 (-1.77- -0.51) |
| PHQ_Score | -3.50 | 1.11 | -3.15 | 94 | $2.2 \times 10^{-3}$ | $3.3 \times 10^{-3}$ | -0.95 (-1.57- -0.34) |
| PILL_Score | -5.29 | 2.28 | -2.32 | 94 | 0.02 | 0.02 | -0.70 (-1.31- -0.09) |
| CMSI_12m_Score | -7.31 | 2.08 | -3.51 | 90 | $7 \times 10^{-4}$ | $2.1 \times 10^{-3}$ | -1.07 (-1.69- -0.44) |
| CMSI_Lftm_Score | -6.42 | 2.17 | -2.95 | 90 | $4 \times 10^{-3}$ | $4.8 \times 10^{-3}$ | -0.90 (-1.52- -0.28) |
| CMSI_SA_Score | -2.96 | 0.92 | -3.21 | 90 | $1.9 \times 10^{-3}$ | $3.3 \times 10^{-3}$ | -0.97 (-1.60 - -0.35) |

SR, Stress Reactivity; SE, standard error; df, degrees of freedom; CMSI_SS_Score, Complex Medical Symptom Inventory Sensory Sensitivity; PHQ_Score, Patient Health Questionnaire without gastrointestinal items; PILL, Pennebaker Inventory of Limbic Languidness; CMSI_12m_Score, Complex Medical Symptom Inventory reporting presence of somatic symptoms for 3 months out of the past year; CMSI_Lftm_Score, Complex Medical Symptom Inventory reporting presence of somatic symptoms over the lifetime; CMSI_SA_Score, Complex Medical Symptom Inventory Somatic Awareness

Supplemental Table 8. Comorbidity Differences between HCs in the Low Compared to the High SR Groups

| **Variables** | **t** | **df** | ***P*** | **q** | **Cohen's d (95% CIs)** |
|---|---|---|---|---|---|
| CMSI_SS_Score | -2.40 | 57 | 0.02 | 0.07 | -0.8 (-1.49--0.12) |
| PHQ_Score | -1.17 | 68 | 0.25 | 0.25 | -0.36 (-0.97-0.26) |
| PILL_Score | -1.93 | 68 | 0.06 | 0.10 | -0.59 (-1.22-0.03) |
| CMSI_12m_Score | -2.08 | 57 | 0.04 | 0.10 | -0.69 (-1.38--0.01) |
| CMSI_Lftm_Score | -1.38 | 57 | 0.17 | 0.20 | -0.46 (-1.14-0.21) |
| CMSI_SA_Score | -1.74 | 57 | 0.09 | 0.12 | -0.58 (-1.26-0.10) |

HC, health control; SR, stress reactivity; df, degrees of freedom; CMSI_SS_Score, Complex Medical Symptom Inventory Sensory Sensitivity; PHQ_Score, Patient Health Questionnaire without gastrointestinal items; PILL, Pennebaker Inventory of Limbic Languidness; CMSI_12m_Score, Complex Medical Symptom Inventory reporting presence of somatic symptoms for 3 months out of the past year; CMSI_Lftm_Score, Complex Medical Symptom Inventory reporting presence of somatic symptoms over the lifetime; CMSI_SA_Score, Complex Medical Symptom Inventory Somatic Awareness.

Supplemental Table 9. Means by High and Low SR Groups in HC's

| Variables | Low SR | | | High SR | | |
|---|---|---|---|---|---|---|
| | *M* | SE | n | *M* | SE | n |
| CMSI_SS_Score | 0.10 | 0.06 | 48 | 0.55 | 0.28 | 11 |
| PHQ_Score | 2.13 | 0.29 | 57 | 2.91 | 0.51 | 13 |
| PILL_Score | 4.23 | 0.54 | 57 | 6.62 | 0.99 | 13 |
| CMSI_12m_Score | 1.06 | 0.42 | 48 | 3.18 | 1.07 | 11 |
| CMSI_Lftm_Score | 1.69 | 0.59 | 48 | 3.55 | 1.08 | 11 |
| CMSI_SA_Score | 0.50 | 0.19 | 48 | 1.27 | 0.41 | 11 |

SR, stress reactivity; HC, healthy controls; CMSI_SS_Score, Complex Medical Symptom Inventory Sensory Sensitivity; PHQ_Score, Patient Health Questionnaire without gastrointestinal items; PILL, Pennebaker Inventory of Limbic Languidness; CMSI_12m_Score, Complex Medical Symptom Inventory reporting presence of somatic symptoms for 3 months out of the past year; CMSI_Lftm_Score, Complex Medical Symptom Inventory reporting presence of somatic symptoms over the lifetime; CMSI_SA_Score, Complex Medical Symptom Inventory Somatic Awareness

Supplemental Table 10. Significant Clusters Showing Lower MD in High Compared to Low SR Groups

| Cluster Number | Tract ROI | Cluster Volume ($\mu L$) | Centroid |
|---|---|---|---|
| **1** | JHU Anterior Limb Internal Capsule (L)<br>JHU Anterior Limb Internal Capsule (R)<br>JHU Posterior Limb Internal Capsule (L)<br>JHU Posterior Limb Internal Capsule (R)<br>JHU Retrolenticular part Internal Capsule (L)<br>JHU Retrolenticular part Internal Capsule (R)<br>JHU External Capsule (L)<br>JHU External Capsule (R)<br>JHU Anterior Corona Radiata (R)<br>JHU Posterior Corona Radiata (R)<br>JHU Superior Corona Radiata (R)<br>JHU Corticospinal Tract (L)<br>JHU Corticospinal Tract (R)<br>JHU Body Corpus Callosum<br>JHU Genu Corpus Callosum<br>JHU Fornix Stria Terminalis (R)<br>JHU Sagittal Stratum (R)<br>JHU Cerebral Peduncle (L)<br>JHU Cerebral Peduncle (R)<br>JHU Cingulum Cingulate Gyrus (R)<br>JHU Superior Fronto-occipital Fasciculus (L)<br>JHU Superior Fronto-occipital Fasciculus (R)<br>JHU Superior Longitudinal Fasciculus (R)<br>JHU Uncinate Fasciculus (R)<br>Thalamus | 32791 | (-42.0 +8.0 -32.0) |
| **2** | JHU Anterior Limb Internal Capsule (L)<br>JHU Body Corpus Callosum<br>JHU Genu Corpus Callosum<br>JHU Cingulum Cingulate Gyrus (L)<br>JHU Superior Corona Radiata (L) | 4537 | (+14.0 -36.0 -18.0) |
| **3** | JHU Retrolenticular part Internal Capsule (R)<br>JHU Posterior Corona Radiata (R)<br>JHU Sagittal Stratum (R)<br>JHU Posterior Thalamic Radiation (R)<br>JHU Superior Longitudinal Fasciculus (R)<br>Thalamus | 2025 | (-38.0 +44.0 -11.0) |
| **4**<br>(+25.0 +13.0 -12.0) | JHU Anterior Limb Internal Capsule (L)<br>JHU Anterior Corona Radiata (L)<br>JHU External Capsule (L) | 1888 | |

SR, stress reactivity; ROI, region of interest; µL, microliter; JHU, Johns Hopkins University; R, right; L, left.

Supplemental Table 11. Low - High SR group Differences in the Resting State Connectivity of the Central Autonomic Network

| DVs | Description | Region name | Network | t | *P* | q | Cohen's d (95% CIs) |
|---|---|---|---|---|---|---|---|
| R_InfCirIns | Inferior segment of the circular sulcus of the insula | Posterior Insula | Homeostatic afferent/sensorimotor | 4.12 | $5.94 \times 10^{-5}$ | $2.00 \times 10^{-3}$ | 0.70 (0.36-1.05) |
| L_InfCirIns | Inferior segment of the circular sulcus of the insula | Posterior Insula | Homeostatic afferent/sensorimotor | 3.56 | $4.89 \times 10^{-4}$ | $1.00 \times 10^{-2}$ | 0.61 (0.26-0.95) |
| R_LoInG_CinS | Long insular gyrus and central sulcus of the insula | Posterior Insula | Homeostatic afferent/sensorimotor | 3.31 | $1.17 \times 10^{-3}$ | $1.60 \times 10^{-2}$ | 0.56 (0.22-0.90) |
| L_Hip | Hippocampus | Hippocampus | Emotional Arousal | 3.08 | $2.45 \times 10^{-3}$ | $2.50 \times 10^{-2}$ | 0.52 (0.18-0.86) |
| R_PosLS | Posterior ramus of the lateral sulcus | Posterior Insula | Homeostatic afferent/sensorimotor | 3.02 | $2.98 \times 10^{-3}$ | $2.50 \times 10^{-2}$ | 0.51 (0.17-0.85) |
| R_SupCirInS | Superior segment of the circular sulcus of the insula | Anterior Insula | Salience | 2.89 | $4.44 \times 10^{-3}$ | $3.10 \times 10^{-2}$ | 0.49 (0.15-0.83) |
| L_SupCirInS | Superior segment of the circular sulcus of the insula | Anterior Insula | Salience | 2.83 | $5.00 \times 10^{-3}$ | $3.20 \times 10^{-2}$ | 0.48 (0.14-0.82) |
| L_Amg | Amygdala | Amygdala | Emotional Arousal | 2.74 | $7.00 \times 10^{-3}$ | $3.60 \times 10^{-2}$ | 0.47 (0.13-0.81) |
| L_LoInG_CinS | Long insular gyrus and central sulcus of the insula | Posterior Insula | Homeostatic afferent/sensorimotor | 2.52 | $1.30 \times 10^{-2}$ | 0.06 | 0.43 (0.09-0.77) |
| R_Amg | Amygdala | Amygdala | Emotional Arousal | 2.30 | $2.30 \times 10^{-2}$ | 0.09 | 0.39 (0.05-0.73) |
| L_AcgG_S | Anterior part of the cingulate gyrus and sulcus(ACC) | Rostaral anterior cingulate | Emotional Arousal | 2.31 | $2.20 \times 10^{-2}$ | 0.09 | 0.39 (0.06-0.73) |
| L_PaHipG | Parahippocampal gyrus | Parahippocampal gyrus | Emotional Arousal | 2.23 | $2.70 \times 10^{-2}$ | 0.09 | 0.38 (0.04-0.72) |
| R_Hip | Hippocampus | Hippocampus | Emotional Arousal | 2.15 | $3.30 \times 10^{-2}$ | 0.10 | 0.37 (0.03-0.70) |

*Supplemental Table 11 Continued*

| | | | | | | | |
|---|---|---|---|---|---|---|---|
| L_PrCun | Precuneus | Precuneus | Default mode | 2.14 | $3.40 \times 10^{-2}$ | 0.10 | 0.36 (0.03-0.70) |
| L_mPFC | medial prefrontal cortex | Medial prefrontal cortex | Default mode | 2.19 | $3.00 \times 10^{-2}$ | 0.10 | 0.37 (0.03-0.71) |
| L_ShoInG | Short insular gyri | Anterior insula | Salience | 1.99 | $4.90 \times 10^{-2}$ | 0.13 | 0.34 (0.01-0.68) |
| R_PBC | parabrachial complex | Parabrachial complex | Homeostatic afferent/sensorimotor | 1.93 | 0.06 | 0.14 | 0.33 (-0.01-0.67) |
| R_PaHipG | Parahippocampal gyrus | Parahippocampal gyrus | Emotional Arousal | 1.87 | 0.06 | 0.14 | 0.32 (-0.02-0.66) |
| R_PrCun | Precuneus | Precuneus | Default mode | 1.87 | 0.06 | 0.14 | 0.32 (-0.02-0.66) |
| MRN | median raphe | Median raphe | Emotional Arousal | 1.86 | 0.07 | 0.14 | 0.32 (-0.02-0.65) |

SR, stress reactivity; DV, dependent variable; SE, standard error; CI, confidence interval; R, right; L, left

Supplemental Table 12. Significant Clusters Showing Higher FA Values in the High Compared to Low SR Group

| Cluster Number | Tract ROI | Cluster Volume (μL) | Centroid |
|---|---|---|---|
| 1 | JHU Anterior Limb Internal Capsule (R)<br>JHU Posterior Limb Internal Capsule (R)<br>JHU Cerebral Peduncle (R)<br>Thalamus | 1292 | (-6.0 +7.0 -14.0) |
| 2 | JHU Posterior Limb Internal Capsule (L)<br>JHU Cerebral Peduncle (L)<br>JHU Fornix Stria Terminalis (L)<br>Thalamus | 1034 | (+16.0 +25.0 -14.0) |
| 3 | JHU Posterior Corona Radiata (L)<br>JHU Posterior Thalamic Radiation (L)<br>JHU Superior Longitudinal Fasciculus (L) | 1004 | (+35.0 +54.0 -3.0) |

SR, stress reactivity; ROI, region of interest; FA, fractional anisotropy; μL, microliter; JHU, Johns Hopkins University; R, right; L, left.
